# A systematic review of the methods used to ensure rigour, transparency and validity in rapid evaluations, rapid appraisals and rapid assessments

**DOI:** 10.1101/2023.12.20.23300284

**Authors:** Sigrún Eyrúnardóttir Clark, Norha Vera San Juan, Thomas Moniz, Rebecca Appleton, Phoebe Barnett, Cecilia Vindrola-Padros

## Abstract

Rapid approaches are essential when resources are limited in research settings and in the real-time evaluation of changing services. Limitations exist in their design and implementation which can lead to a reduced level of trust in findings. This study identified the methods used across rapid evaluations, appraisals and assessments to improve rigour, transparency and validity. Four scientific databases and one search engine were searched between 11-16th August 2022. Screening led to the inclusion of 169 articles that provided a much-needed repository of methods that can be used during the design and implementation of rapid studies to improve rigour. We suggest that this repository of methods inform the development of transparent reporting standards for future rapid evaluations, appraisals and assessments.

## Introduction

There is an ever growing need to conduct evaluations and research rapidly to deliver findings in a timely way so these can be used to inform decision-making processes (Beebe, 1995; Johnson & Vindrola-Padros, 2017; McNall & Foster-Fishman, 2007). While prolonged in-depth approaches are most appropriate in some occasions, rapid approaches are vital in contexts where time and resources are limited such as humanitarian crises or complex health emergencies, and real-time evaluation of changing programmes and services (Anker et al., 1993; Beebe, 1995; Gawaya et al., 2022; Holdsworth et al., 2020; Johnson & Vindrola-Padros, 2017; McNall & Foster-Fishman, 2007). While rapid approaches are the most appropriate to produce timely findings when they are needed most, limitations can exist in their design and implementation.

There is a wealth of research on the different types of studies that can be classified as rapid approaches. Some of these studies have included Rapid Evaluation Methods (REM); Rapid Cycle Evaluations (RCE); Real-Time Evaluations (RTE); Rapid Feedback Evaluations (RFE); Rapid Appraisals; Rapid Participatory Appraisals; Rapid Rural Appraisals; Participatory Rural Appraisals; Rapid Assessment Methodologies (RAM); Rapid Assessment Procedures or Processes (RAP); Rapid Assessment and Response (RAR); Rapid Assessment Response and Evaluations (RARE); Rapid Assessment Procedure Informed Clinical Ethnography (RAPICE); Rapid Ethnographic Assessments (REA); Rapid Ethnographic Appraisals; and Rapid Ethnography (McNall & Foster-Fishman, 2007; Norman et al., 2022; Vindrola-Padros, 2021; Vindrola-Padros et al., 2020). Rapid research and rapid evaluation are often used interchangeably, however differences between the two exist. Rapid research commonly refers to findings that are delivered in the context of time and resource constraints, and rapid evaluation commonly refers to the delivery of timely evidence to inform decision making and service delivery (Vindrola-Padros, 2021). Key features of the study designs listed above that our team has compiled in previous work include: studies being conducted within a short timeline (anywhere from a few weeks to a few months); a participatory approach, whereby members from the community of interest to the study are involved in the research or evaluation process; team-based approaches as a group can work collaboratively to produce findings in a shorter time period than one researcher/evaluator would be able to; and iterative data collection and analysis as analysis often begins in the field to identify emerging findings as data are still being collected (Vindrola-Padros, 2021; Vindrola-Padros et al., 2020).

Authors have previously identified limitations in these forms of rapid research, for example, not having the time or resources to reach a representative group of study participants; not being able to carry out in-depth data collection or analysis; atheoretical studies and the failure to include critical perspectives in the research; and lack of clarity in the reporting of the methods used and changes made throughout the study (Beebe, 1995; Fitch et al., 2004; Johnson & Vindrola-Padros, 2017; McMullen et al., 2011; McNall & Foster-Fishman, 2007; Norman et al., 2022; Vindrola-Padros, 2021; Vindrola-Padros & Vindrola-Padros, 2018). Not taking steps to minimise these limitations may ultimately lead to a reduced level of trust in the research that has been conducted (Beebe, 1995; McNall & Foster-Fishman, 2007; Vindrola-Padros, 2021). Similarly to rapid research, limitations exist within rapid evaluations, which include limited engagement with evaluation theory; limited depth and breadth of evidence preventing the ability to effectively inform decision making; limited resourcing to fund the time needed for evaluation teams; finally, many rapid evaluations have failed to share information on their dissemination plans (Vindrola-Padros, 2021).

There has been a call to improve the transparency, validity and quality of rapid research and rapid evaluation to improve the trustworthiness and replicability of these studies (McNall & Foster-Fishman, 2007; Vindrola-Padros, 2021). We sought to address this gap in the literature by conducting a systematic review of the methods used to ensure rigour, transparency and validity in rapid evaluation, appraisal and assessment studies.

## Methods

The systematic review was guided by the Preferred Reporting Items for Systematic Reviews and Meta-Analyses (PRISMA) 2020 statement (Page et al., 2021). A protocol outlining the project steps was accepted by PROSPERO prior to conducting the research in August 2022 (CRD42022341825).

### 1. Search strategy

The search strategy for the review included both keyword and subject heading searches across four scientific literature databases: MEDLINE, Embase, Healthcare Management Information Consortium (HMIC), Cumulative Index of Nursing and Allied Health Literature (CINAHL) Plus, and the open search engine Google Scholar. Search words were related to ‘rapid evaluation’, ‘rapid appraisal’ or ‘rapid assessment’. The searches were conducted between 11–16th August 2022. A detailed outline of the search strategy can be found in Appendix 1.

### 2. Eligibility criteria

Criteria for including studies consisted of (1) study methods being rapid evaluations, rapid appraisals, rapid assessments, or any variation of these that were listed in the introduction of this paper; (2) the study having been completed within 6 months (in keeping with rapid approaches) (Vindrola-Padros, 2021); and (3) the article including sufficient information on the methods used to ensure rapidity, transparency, validity and rigour. There were no restrictions on publication date, language or study design in terms of being qualitative, quantitative or mixed methods. Despite conducting the majority of the searches on health-related databases, we did not specify our inclusion criteria to health-related fields.

### 3. Selection process

The search results were imported into EndNote for de-duplication (Gotschall, 2021), and then to Rayyan for further de-duplication and screening (Ouzzani et al., 2016). The title and abstract screening based on the eligibility criteria was split between three researchers due to the large number of search results. The three researchers then cross-checked 10% of each other’s exclusions and discussed any disagreements of exclusion decisions until a consensus on the decision was reached. The inclusion criteria were then refined to ensure all reviewers had the same understanding before the remaining screening was conducted. Following this, one researcher progressed the included articles to full text screening against the eligibility criteria, using Microsoft Excel to report the screening decision. Any articles identified in non-English languages were translated using Google Translate. The principal investigator then cross-checked 10% of the excluded articles.

### 4. Data extraction and quality assessment

A comprehensive list of data extraction categories was developed based on the research question and pre-specified outcomes of interest. These categories were further refined to ensure information was captured appropriately following screening. Data extraction, quality assessments and full text screening was carried out in parallel by one researcher, recording information on Microsoft Excel. The final data extraction categories included: study details (location of study; study design; study duration; general methods used throughout) and the methods used across the different stages of the rapid studies (as shown in Appendix 2) such as during study design; data collection; data analysis; result interpretation; and dissemination. We also extracted any method limitations outlined in papers.

The Mixed Methods Appraisal Tool (MMAT) (Hong et al., 2018) was used to assess the quality of the included articles because of the heterogenous range of study designs. No articles were excluded based on their MMAT scores.

### 5. Data synthesis

Textual narrative synthesis and a summary of content was conducted to summarise the study characteristics and key findings across the literature (Lucas et al., 2007). We then used the rigorous and accelerated data reduction technique (RADaR) to reduce the data within our findings (Watkins, 2017).

## Results

### 1. Study selection

The search returned 33,144 results, of which 20,922 articles remained following duplicate removal. Of these, 577 articles were identified as potentially relevant and screened at full text. Overall, 169 articles were included in the review. Figure 1 shows the flow of studies through the review process based on PRISMA guidelines (Page et al., 2021), and the full list of included references can be found in Appendix 3.

**Figure 1.**
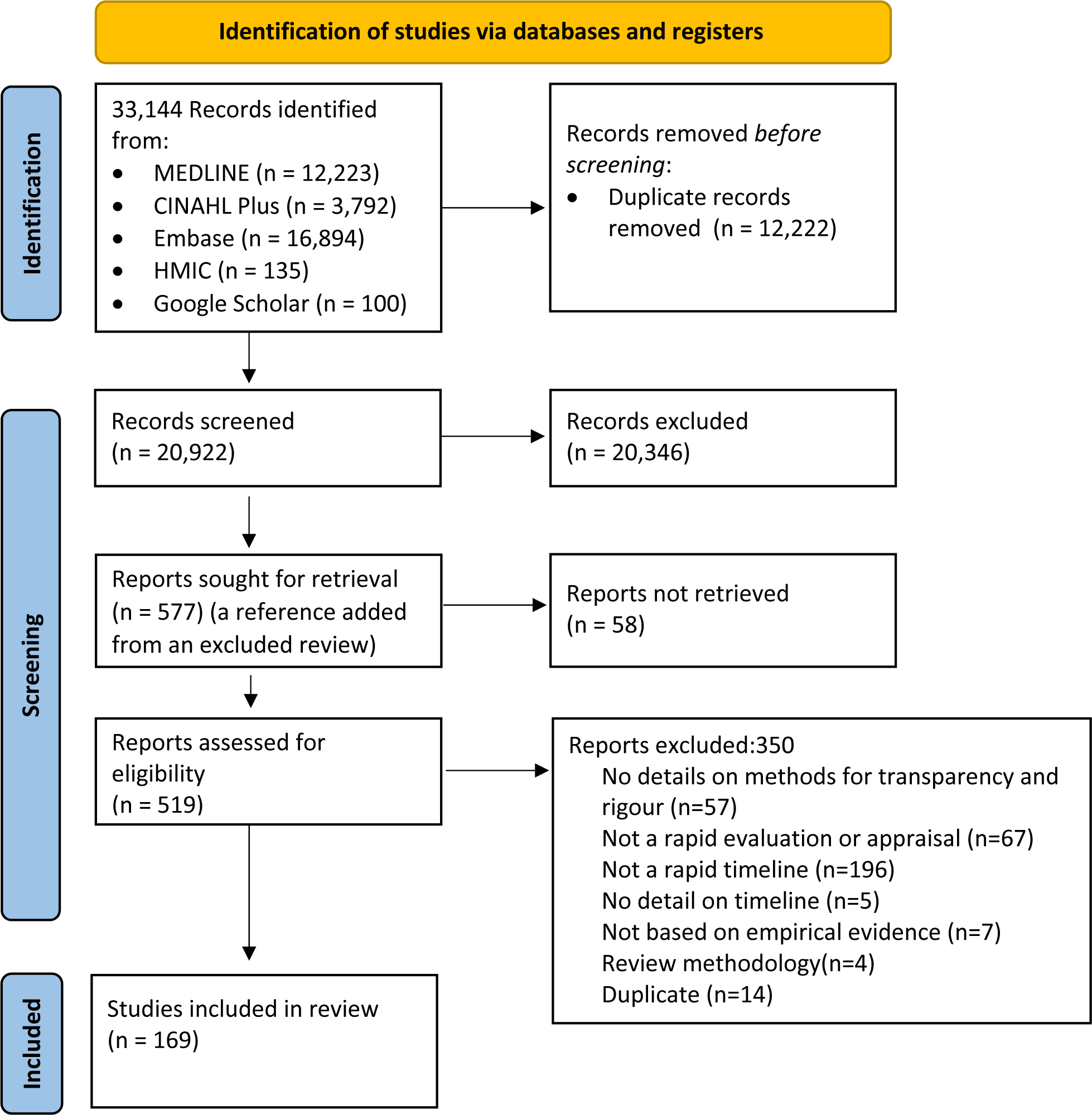
**PRISMA 2020 Flow Diagram of the Selection Process**

The reasons for excluding articles at full text screening can be found in Figure 1. This included articles not sharing enough detail on the methods used to ensure transparency and rigour; the study not fitting under a variation of rapid evaluation or appraisal; the study not being conducted to a rapid timeline (within 6 months); no detail on the study timeline; the study not being based on empirical evidence; the study using review methodology; or the study was a duplicate of a previously screened item.

### 2. Study characteristics

A detailed summary of the characteristics and themes of all the studies included in the review can be found in Appendix 2.

Most of the included studies were conducted in the USA (n=35), followed by the UK (n=12), South Africa (n=9), Uganda (n=8) and Kenya (n=7), more details on study location can be found in Appendix 2. Rapid assessments were the most common form of rapid studies, followed by rapid appraisals and rapid evaluations, more detail can be found in Table 1 below. A large majority of the rapid approaches were conducted within qualitative study designs.

**Table 1.** Summary of the Study Designs of Included Publications.

| Characteristics | Number of studies |
| --- | --- |
| **Type of study design** |  |
| Qualitative | 110 |
| Quantitative | 21 |
| Mixed methods | 38 |
| **Type of rapid study** |  |
| Rapid evaluation | 13 |
| Rapid appraisal | 31 |
| Rapid rural appraisal | 3 |
| Participatory rapid appraisal | 3 |
| Participatory rural appraisal | 9 |
| Participatory needs assessment | 1 |
| Rapid assessment | 88 |
| Rapid assessment response | 6 |
| Rapid assessment response and evaluation | 2 |
| Rapid rural assessment | 1 |
| Peer ethnography | 1 |
| Rapid assessment procedure informed clinical ethnography | 1 |
| Rapid ethnographic assessment | 2 |
| Rapid ethnographic evaluation | 1 |
| Participatory evaluation | 1 |
| Evaluation | 4 |
| Rapid analysis | 2 |

### 3. Quality assessment

The MMAT scores of each study can be found in Appendix 2. The studies ranged from low quality with a score of 0/5 (n=3) and 1/5 (n=32) to medium quality with a score of 2/5 (n=38) and 3/5 (n=25), to high quality with a score of 4/5 (n=61) and 5/5 (n=10).

### 4. Characteristics of rapid studies that ensure rigour and/or transparency

Appendix 2 reports the methods used by each included study, which are also summarised below.

### 4.1 Frameworks, models and reporting guidelines

There were 29 studies that discussed using frameworks, models, theories or existing methodologies to guide their study design. The most common frameworks and models that were cited were the Consolidated Framework for Implementation Research (n=3) and the Health Belief Model (n=3). The most common methodologies that were used to guide the studies were the International Rapid Assessment Response and Evaluation (I-RARE)/RARE methodology (n=4) and the Rapid Assessment Procedure-Informed Clinical Ethnography (RAPICE) methodology (n=3).

There were only six studies that followed reporting guidelines. The Consolidated criteria for reporting qualitative research (COREQ) guidelines were the most commonly cited (n=4).

### 4.2 Study design

In terms of designing and setting up the study a number of approaches were reported that enabled transparency, rigour and rapidity of studies. There were 41 publications that specifically mentioned following a study protocol or proposal, and six articles that discussed working with a steering group to guide the study design.

Eight articles highlighted they did not need to subject their study to Institutional Review Committees as the projects were classified as non-research and instead classified as service evaluations, or they worked with online anonymised surveys. One study received informal approvals and comments from the technical team of an Institutional Review Committee. Two studies discussed sharing research plans with the local organisations and areas concerned with the evaluations.

The majority of the included articles (n=91) obtained informed consent from participants ahead of data collection in the form of verbal, written and assumed consent (based on voluntary participation in online surveys). There were two studies that shared that consent was not obtained due to examination of services or observational designs, however it was made clear to participants observations were taking place.

A large majority of the publications discussed using sampling methodologies to either increase rapidity or to improve relevance of characteristics of the sample. The main form of sampling discussed in 84 of the studies was the use of non-probability sampling in the form of convenience sampling, purposive sampling or variations of snowball sampling. Fourteen of the studies discussed how sample size was determined based on when data saturation was reached. Three articles discussed methods used to minimise biases in sampling by searching for alternative or disconfirming cases, deviating from arranged locations to try to reach unsensitized groups, and using active, sequential recruitment.

In terms of recruitment, 22 of the articles discussed having support from local key informants, local researchers/evaluators, local networks and organisations to build trust with potential study participants and to advertise the study. Sixteen of the articles discussed recruiting participants via email or through social networking via WhatsApp or social media.

Some of the articles discussed using incentivisation to improve recruitment. Twenty-three of the articles discussed using monetary incentives either in the form of honorarium or in the form of vouchers. Six of the articles discussed using food and drinks as incentives, and four of the articles discussed health incentivisation through referring participants onto support services and providing condoms. There were five articles that specifically mentioned that their study participants were not incentivised or compensated for their participation.

### 4.3 Data collection

Methods of data collection that improved transparency, validity and rapidity of the studies included piloting methods, using numerous methods for data collection, collecting data in local languages, working with large experienced multidisciplinary teams of researchers/evaluators, conducting team meetings, using participatory methods and recording, transcribing and developing field notes.

There were 26 publications that discussed piloting and reviewing data collection tools to ensure cultural and face validity. The majority of the publications (n=82) also discussed using numerous methods of data collection in the form of interviews, focus groups, surveys and observations for subsequent triangulation.

A large number (n=66) of the studies conducted research or evaluations in local languages working with translators, local researchers/evaluators and with researchers with lived experience. There were nine studies that discussed how this helped to facilitate trust and strong validity of the cultural and traditional knowledge.

The majority of the studies (n=87) discussed the composition and experience of the evaluation or research team. They discussed whether they worked in large multidisciplinary teams with individuals with diverse experiences and previous experiences in research or evaluation. They also shared whether researchers/evaluators had undergone training or were under the supervision of a senior team member. Twenty-two of the studies discussed conducting team meetings during data collection to ensure good data quality, validity and representativeness of samples and to modify any processes or materials.

There were 14 studies that discussed using participatory methods for data collection such as participating in existing community meetings and using mind maps, matrix ranking and other techniques to elicit information for action, and to generate group discussions allowing for room to correct any misinterpretations.

Twenty-three of the studies discussed using technology for data collection, by conducting interviews and focus groups over video conferencing software, over the phone and over social media platforms like WhatsApp. They also discussed conducting surveys through online platforms and using data from online Twitter forums. The majority of the studies (n=84) also discussed recording and transcribing interviews and reviewing any field notes either in place of transcripts or to enable early analysis of data ahead of availability of transcripts.

### 4.4 Data analysis

Many of the qualitative studies (n=65) discussed using forms of qualitative content analysis including thematic analysis or the Framework approach both using stepwise approaches to data familiarisation, coding, generating themes and reviewing themes, with the latter method having a focus on using matrices or tables to aid analysis. There were 12 papers that discussed using the ‘coding consensus, co-occurrence, and comparison’ approach or the Grounded Theory approach to openly code transcripts and use the constant comparative method to identify connections between the data. There were 11 studies that discussed specifically using rapid analysis techniques such as rapid assessment procedure sheets. One study discussed using the RADaR technique to reduce data and two studies discussed using mind-mapping techniques to help to visualise the data.

Iterative data collection and data analysis was mentioned across 32 studies. They reported that analysing data whilst data collection was ongoing allowed teams to identify when data saturation had been met, to re-shape data collection tools based on emerging findings and to develop initial codebooks for subsequent in-depth analysis. The method of data triangulation was also commonly used across 58 publications. The methods included triangulating data from different subgroups of participants that data were collected from, data from different data collection methods such as surveys and interviews, and data from existing secondary data sources such as historical records and existing statistical data.

There were 42 studies that shared their team had meetings to compare any emerging findings, discuss any discrepancies in analysis and to reach a consensus on the analysis. Two of the studies discussed using field notes of the team members analysis as an audit trail. Twenty-two of the studies discussed using quality assurance methods during analysis to assess consistency in analysis across team members and to assess the quality of the analysed data.

### 4.5 Result interpretation

Some of the studies went on to discuss how teams interpreted study results, with four of the studies sharing that they conducted team meetings to discuss findings and reach consensus on how best to interpret them. Six of the studies shared that their team underwent a form of reflexive thinking to reflect on their practice throughout the study and how their personal characteristics may have affected their interpretations of the results. There were three studies that shared that they had a peer-review team that reviewed the research/evaluation teams’ interpretations and the draft reports to identify any potential biases.

Many of the studies (n=27) discussed using member checking to validate the findings from the study with the study participants themselves or with key informants from the study. Seven studies also shared that advisory groups separate to the evaluation/research team supported with interpreting interim and initial findings.

### 4.6 Dissemination

Using iterative dissemination of findings whilst studies were ongoing was discussed in three studies to rapidly share emerging findings with key stakeholders. The studies also shared that this process allowed feedback loops to be created between stakeholders implementing findings and the evaluation/research team, enabling the evaluation of studies or programs to take place as they are being implemented.

## 5. Limitations of the methods used in rapid studies

The final part of this analysis included a summary of the limitations the authors identified in the rapid studies. The most frequent limitation were the short timelines that made it difficult to enable in-depth study planning, data collection, analysis, interpretation and to disseminate findings in a meaningful way. These limitations have been summarised in Table 2.

**Table 2.** The Limitations of the Methods Used in Rapid Studies.

| **Phase of Study** | **Limitations** |
| --- | --- |
| Study design | • Short timelines prevented in-depth study planning.<br>• Small sample sizes limited generalisability and representativeness of findings.<br>• Non-probability sampling introduced selection bias.<br>• Difficulties setting up working groups without existing connections. |
| Data collection | • Short timelines prevented in-depth data collection.<br>• Short timelines prevented staff training and pre-testing collection instruments.<br>• Mind maps introduced researcher bias.<br>• Inability to audio record interviews.<br>• Online surveys led to selection bias and ambiguity on non-response rate and sampling frames.<br>• Non-face-to-face interviews may have prevented open responses.<br>• Conducting data collection in non-native language affected level of detail of response.<br>• Loss of contextual validity during translations.<br>• Response bias due to type of researcher and location of data collection. |
| Data analysis | • Short timelines prevented in-depth data analysis.<br>• Inability to corroborate findings with other sources.<br>• Interim analysis preventing attribution of findings to an instrument or participant.<br>• Over or underrepresenting themes during analysis.<br>• Difficulties auditing analysis as decisions made during discussions. |
| Result interpretation and team dynamics | • Researcher's experiences affecting interpretations of findings.<br>• Wellbeing and safety risks to team members.<br>• Under-estimating resource requirements. |
| Result dissemination | • Short timelines prevented meaningful sharing of findings. |

### 5.1 Limitations encountered during study design

In terms of study design, the majority of publications identified having small sample sizes as a limitation, which led to limited generalisability and representativeness of findings. Also mentioned was the introduction of selection bias of participants and limited generalisability of findings because of non-probability sampling. Some of the publications also shared they found it difficult to set up advisory groups and working groups if connections with the groups were not already in place.

### 5.2 Limitations encountered during data collection

Some of the publications discussed not having enough time to train staff in the methods they had planned to use, or to pre-test the data collection instruments, which affected consistency in data collection between team members. Using mind maps as a form of data collection was identified as a limitation as it did not allow enough time for participants to reflect on the data they had shared, and it could be heavily impacted by evaluator/researcher bias. Some of the studies were unable to audio record interviews due to privacy concerns which affected the accuracy of findings. Using online data collection made it difficult for some studies to determine the non-response rate, sampling frame, and could have led to selection bias of participants that could access and use online technology. Conducting interviews over the phone or through teleconferencing software was thought to affect how open respondents were to questions. Some studies identified that having team members that did not speak the local language affected interpretations of findings and that conducting research or evaluations in a foreign language to the study participants, affected how detailed responses were. The studies also identified that some contextual validity was lost following translation. It was frequently reported that a form of response bias may have occurred based on participants feeling uncomfortable around the type of researcher/evaluator for example based on their gender, age or whether a local researcher/evaluator who they may know was collecting their data. Similarly, response bias was flagged as a potential limitation depending on the location of data collection, for instance in a workplace.

### 5.3 Limitations encountered during data analysis

Some of the studies mentioned the limitation of being unable to triangulate findings with other sources to corroborate self-reported responses from participants that may have been prone to recall or response bias. Conducting interim analysis was also identified as a limitation as it made it difficult to attribute findings to a particular collection instrument or participant. The issue of over-representing or under-representing themes during analysis was also reported, as were the difficulties in auditing analysis when many decisions around coding had taken place during group discussions rather than during physical coding.

### 5.4 Limitations encountered during the result interpretation and within team dynamics

Aspects of the team were discussed within the studies as limitations, including team member characteristics and experiences affecting interpretations of findings. Additionally, the difficulty and safety risks of the team when accessing study sites were identified as a limitation, as was the impact of stressful rapid timelines on team member wellbeing. Finally, it was reported in some of the studies that they had under-estimated their resource requirements so did not have enough team members to plan or conduct the study.

## Discussion

The aim of this review was to contribute to the field of rapid appraisals, evaluations and assessments, and identify the methods that have been used by evaluators and researchers to maintain the quality of the studies in the context of time pressures. The review is a response to current debates in the literature on the quality and rigour of rapid studies, where short timeframes are often associated with ‘quick and dirty’ exercises, rushed processes or studies where evaluators and researchers might be ‘cutting corners’ (Cupit et al., 2018; Vindrola-Padros, 2021; Vindrola-Padros & Vindrola-Padros, 2018).

We identified a wide range of methods used to increase the speed of evaluations and research, particularly during stages of data collection and analysis. This included using non-probability sampling; using existing networks to support with recruitment; incentivisation for recruitment; team-based approaches for data collection and analysis; technology to support with data collection; rapid analysis techniques; and an iterative process of data collection, analysis and dissemination. We found that evaluators/researchers also used methods to ensure transparency, quality and rigour whilst conducting rapid studies, such as the use of reporting guidelines, integrating conceptual frameworks to guide data collection and interpretation, developing and adhering to protocols, working with advisory groups to guide the project, and gaining regulatory approvals and consent from study participants. Other methods included piloting data collection methods; conducting evaluations or research in local languages; working as multidisciplinary teams with relevant training in the methods used; undergoing team discussions on discrepancies in methods used; using participatory methods to collect data; triangulating data sources; using member checking and peer-review to confirm interpretations; and reflexive thinking to highlight how evaluator/researcher experience may affect interpretations.

The limitations outlined in the articles included in the review corroborated limitations previously discussed in the literature on rapid evaluation and rapid research. Short timelines can hinder the ability for in-depth study planning, data collection, analysis, interpretation and dissemination (Beebe, 1995; Fitch et al., 2004; Johnson & Vindrola-Padros, 2017; McMullen et al., 2011; McNall & Foster-Fishman, 2007; Vindrola-Padros, 2021; Vindrola-Padros & Vindrola-Padros, 2018). Many of the included studies and existing literature have flagged issues of small sample sizes and the non-random sampling methodologies of rapid studies, that limit the generalisability and representativeness of findings (Ahoua et al., 2006; Akello et al., 2007; Anastasaki et al., 2022; Brittain et al., 2019; Butler et al., 2021; Ezard et al., 2011; Hammond et al., 2022; Johnson & Vindrola-Padros, 2017; Morojele et al., 2006; Negandhi et al., 2017; Nemser et al., 2018; Norman et al., 2022; Sy et al., 2020; Theiss-Nyland et al., 2016; Vindrola-Padros, 2021; Vindrola-Padros & Vindrola-Padros, 2018). Other authors have also highlighted the potential atheoretical and instrumental nature of rapid approaches (Cupit et al., 2018). The articles included in the systematic review highlighted strategies that can be used to address common challenges such as lack of inclusion and the reliance on easily accessible sites and study participants. Purposive sampling, iterative methods of recruitment and increasing recruitment time were used to reduce the risk of excluding participants with underrepresented characteristics (Cohn et al., 2021; Jumbe et al., 2021).

Inconsistency in data collection methods and data analysis methods, due to not having the time to train team members or having the time for supervisors to attend all data collection to ensure alignment across team members, were identified as challenges in the rapid studies (Ash et al., 2016; Bayleyegn et al., 2006). Methods such as team debriefings at the end of each day of data collection or during data analysis were used frequently across the included literature for team members to share feedback with each other to ensure consistency with the methods they have and continue to use.

Several authors also reflected on the fact that short timelines often limited the detailed planning of data collection instruments, stopped evaluators/researchers from understanding the context which could lead to sharing misleading information, and inhibited team members from going in depth to ask certain questions to immerse themselves into data collection (Ezard et al., 2011; Seidel et al., 2018; Tindana et al., 2012). Strategies to overcome these challenges included using member checking to share preliminary findings with the participants themselves or with members of their community, as a way to corroborate the analysis and interpretations that had been conducted. Additionally, triangulating findings with other data sources such as documents or surveys about the area of interest was also used to verify findings and interpretations.

Another key limitation identified previously and within this review has been the impact of rapid approaches on the evaluator/researcher wellbeing in terms of their safety whilst in the field and the impact of short timelines on the stress they experience (Ash et al., 2008; McMullen et al., 2011; Rankl et al., 2021; Vindrola-Padros, 2020). The pressure to collect information in condensed timelines can cause stress and exhaustion to the team. Working in cohesive teams has been identified as a way to relieve some of the pressure as the team can share the load to deliver the findings faster, and support each other emotionally (Ash et al., 2008; McMullen et al., 2011; Rankl et al., 2021; Vindrola-Padros, 2020).

The strengths of this review include the wealth of incorporated studies (n=169) that have been published over a relatively long period of time (1993 to 2022). The review itself is broad as the topics included focus on health; disaster management; sociocultural-political areas; organisational change; and agriculture and farming. However, this too serves as a limitation within the methodology of the review. The inclusion criteria were kept broad to include topics not only specific to health, however the searches were only conducted across four health related databases, and one broad search engine. This may have biased the topics of the included articles towards topics on health. This issue could be overcome by searching alternative databases and working with librarians with specialities in other fields. The type of literature that was included in this review were articles that had been published in peer-review journals, which meant many rapid evaluation, appraisals or assessments that were not published, were not included, which serves as a limitation. However, the unpublished literature may have been less relevant as these articles may have been less likely to share details on the methods used to ensure rigour.

Many publications were excluded because they did not define the duration of their study, and may therefore have been excluded when they were in fact conducted within six months. This, however, is mainly a limitation with the studies of interest and has been cited previously (Johnson & Vindrola-Padros, 2017; Vindrola-Padros & Vindrola-Padros, 2018). It is therefore recommended that future rapid studies define their study duration clearly, so conclusions can be made around whether or not others consider their timeline rapid. We excluded many articles because they did not extensively report on the methods used to ensure rapidity or transparency. This highlights an issue with the current literature and is a recommendation that future rapid studies report in greater detail the methods they have used (Johnson & Vindrola-Padros, 2017; Vindrola-Padros, 2021; Vindrola-Padros & Vindrola-Padros, 2018). Only six of the included articles in the review discussed following reporting guidelines to guide their study design. This finding has been similar to existing literature that has flagged limited reporting guidelines exist in the field of rapid evaluation and rapid research (Vindrola-Padros, 2021). Highlighting a gap in this field and an area for future development that could enable transparent reporting of methods that improve the validity and rigour of rapid evaluations, appraisals and assessments.

## Conclusion

Our systematic review has contributed to the field by sharing the methods that have been used to increase rapidity whilst maintaining transparency and rigour across rapid evaluations, appraisals and assessments that have been conducted within six months. We have identified the limitations of some of these methodologies and rapid dynamics.

There will continue to be a need to conduct rapid evaluations and rapid research, as populations are likely to face future pandemics and humanitarian disasters; changes in governmental powers and polices; and changes to health and public sector organisation, all of which would require the timely delivery of findings. It is, therefore, necessary that future rapid evaluations and appraisals use methods to improve the trustworthiness and rigour of findings and clearly report these methods to enable future evaluators and researchers to learn from. Our research team plans to develop reporting standards based on the approaches identified in this review, that will facilitate the future transparent reporting and uptake of methods that ensure rigour and validity of rapid evaluations, appraisals and assessments.

## Data Availability

All data produced in the present study are available upon reasonable request to the authors

## Notes

### Competing Interest Statement

The authors have declared no competing interest.

### Funding Statement

This work was supported by the UKRI MRC Better Methods Better Research grant [grant number: MR/W020769/1].

