## Appendices for "A systematic review of the methods used to ensure rigour, transparency and validity in rapid evaluations, rapid appraisals and rapid assessments"

**Appendix 1 – Search criteria**

<u>Ovid MEDLINE(R) (searched on 11 August 2022)</u>
1 Rapid evaluation.ti,ab. 1764
2 Real time evaluation.ti,ab. 563
3 Quick evaluation.ti,ab. 178
4 Short evaluation.ti,ab. 70
5 Instant evaluation.ti,ab. 12
6 Rapid cycle evaluation.ti,ab. 8
7 ((Fast or faster) adj (assessment? or report* or qualitative research or research or studies or evaluation? or appraisal? or approach* or response brief? or response program*)).ti,ab. 1231
8 ((Limit* or short* or tight or efficient or pragmatic) adj2 (deadline? or time or timescale or timeline) adj2 (assessment? or report? or qualitative research or research or studies or study or evaluation? or appraisal? or approach* or response brief? or response program*)).ti,ab. 1307
9 ((Speed or speedy) adj (assessments or reports or qualitative research or research or studies or evaluations or appraisals or approaches or response briefs or response program*)).ti,ab. 110
10 (Abbreviated adj (assessments or reports or qualitative research or research or studies or evaluations or appraisals or approaches or response briefs or response program*)).ti,ab. 16
11 (Accelerated adj (assessments or reports or qualitative research or research or studies or evaluations or appraisals or approaches or response briefs or response program*)).ti,ab. 211
12 (Brief adj (assessments or reports or qualitative research or research or studies or evaluations or appraisals or approaches or response program*)).ti,ab. 603
13 (Constrained by time and (assessments or reports or qualitative research or research or studies or evaluations or appraisals or approaches or response briefs or response program*)).ti,ab. 16
14 (Expedited adj (assessments or reports or qualitative research or research or studies or evaluations or appraisals or approaches or response briefs or response program*)).ti,ab. 23
15 (Express adj (assessments or reports or qualitative research or research or studies or evaluations or appraisals or approaches or response briefs or response program*)).ti,ab. 8
16 (Immediate adj (qualitative research or research or studies or evaluations or appraisals or approaches or response briefs or response program*)).ti,ab. 73
17 ((Quick adj2 dirty) and (assessments or reports or qualitative research or research or studies or evaluations or appraisals or approaches or response briefs or response program*)).ti,ab. 19
18 (Quick* adj (assessments or reports or qualitative research or research or studies or evaluations or appraisals or approaches or response briefs or response program*)).ti,ab. 84
19 (Stream-line* adj (assessments or reports or qualitative research or research or studies or evaluations or appraisals or approaches or response briefs or response program*)).ti,ab. 1
20 (Streamline* adj (assessments or reports or qualitative research or research or studies or evaluations or appraisals or approaches or response briefs or response program*)).ti,ab. 79
21 (Timeliness adj6 (assessments or reports or qualitative research or research or studies or evaluations or appraisals or approaches or response briefs or response program*)).ti,ab. 262
22 (Timely adj (assessments or reports or qualitative research or research or studies or evaluations or appraisals or approaches or response briefs or response program*)).ti,ab. 209
23 ((Fast or faster) adj (appraisal or approach or study or assessment or report)).ti,ab. 715
24 ((Limit* or short* or tight or efficient or pragmatic) adj2 (deadline* or time or timescale or timeline) adj2 (appraisal or approach or study or assessment or report)).ti,ab. 843
25 ((Speed or speedy) adj (appraisal or approach or study or assessment or report)).ti,ab. 144
26 (Abbreviated adj (appraisal or approach or study or assessment or report)).ti,ab. 52
27 (Accelerated adj (appraisal or approach or study or assessment or report)).ti,ab. 112
28 (Constrained by time and (appraisal or approach or study or assessment or report)).ti,ab. 29
29 (Expedited adj (appraisal or approach or study or assessment or report)).ti,ab. 32
30 (Express adj (appraisal or approach or study or assessment or report)).ti,ab. 61

31 (Immediate adj (appraisal or approach or study or assessment or report)).ti,ab. 432
32 ((Quick adj2 dirty) and (appraisal or approach or study or assessment or report)).ti,ab. 26
33 (Quick* adj (appraisal or approach or study or assessment or report)).ti,ab. 580
34 (Rapid adj (appraisal or approach or study or assessment or report)).ti,ab. 5194
35 (Stream-line* adj (appraisal or approach or study or assessment or report)).ti,ab. 4
36 (Streamline* adj (appraisal or approach or study or assessment or report)).ti,ab. 324
37 (Timeliness adj6 (appraisal or approach or study or assessment or report)).ti,ab. 369
38 (Timely adj (appraisal or approach or study or assessment or report)).ti,ab. 501
39 Participatory rural appraisal.ti,ab. 132
40 Rapid rural appraisal.ti,ab. 29
41 1 or 2 or 3 or 4 or 5 or 6 or 7 or 8 or 9 or 10 or 11 or 12 or 13 or 14 or 15 or 16 or 17 or 18 or 19 or 20 or 21 or 22 or 23 or 24 or 25 or 26 or 27 or 28 or 29 or 30 or 31 or 32 or 33 or 34 or 35 or 36 or 37 or 38 or 39 or 40 14763
42 limit 41 to yr="2002 -Current" 12223

Ovid Embase (searched on 11 August 2022)
1 Rapid evaluation.ti,ab. 2198
2 Real time evaluation.ti,ab. 812
3 Quick evaluation.ti,ab. 256
4 Short evaluation.ti,ab. 110
5 Instant evaluation.ti,ab. 15
6 Rapid cycle evaluation.ti,ab. 13
7 ((Fast or faster) adj (assessment? or report* or qualitative research or research or studies or evaluation? or appraisal? or approach* or response brief? or response program*)).ti,ab. 1509
8 ((Limit* or short* or tight or efficient or pragmatic) adj2 (deadline? or time or timescale or timeline) adj2 (assessment? or report? or qualitative research or research or studies or study or evaluation? or appraisal? or approach* or response brief? or response program*)).ti,ab. 1788
9 ((Speed or speedy) adj (assessments or reports or qualitative research or research or studies or evaluations or appraisals or approaches or response briefs or response program*)).ti,ab. 146
10 (Abbreviated adj (assessments or reports or qualitative research or research or studies or evaluations or appraisals or approaches or response briefs or response program*)).ti,ab. 23
11 (Accelerated adj (assessments or reports or qualitative research or research or studies or evaluations or appraisals or approaches or response briefs or response program*)).ti,ab. 270
12 (Brief adj (assessments or reports or qualitative research or research or studies or evaluations or appraisals or approaches or response program*)).ti,ab. 641
13 (Constrained by time and (assessments or reports or qualitative research or research or studies or evaluations or appraisals or approaches or response briefs or response program*)).ti,ab. 18
14 (Expedited adj (assessments or reports or qualitative research or research or studies or evaluations or appraisals or approaches or response briefs or response program*)).ti,ab. 36
15 (Express adj (assessments or reports or qualitative research or research or studies or evaluations or appraisals or approaches or response briefs or response program*)).ti,ab. 11
16 (Immediate adj (qualitative research or research or studies or evaluations or appraisals or approaches or response briefs or response program*)).ti,ab. 86
17 ((Quick adj2 dirty) and (assessments or reports or qualitative research or research or studies or evaluations or appraisals or approaches or response briefs or response program*)).ti,ab. 20
18 (Quick* adj (assessments or reports or qualitative research or research or studies or evaluations or appraisals or approaches or response briefs or response program*)).ti,ab. 102
19 (Stream-line* adj (assessments or reports or qualitative research or research or studies or evaluations or appraisals or approaches or response briefs or response program*)).ti,ab. 1
20 (Streamline* adj (assessments or reports or qualitative research or research or studies or evaluations or appraisals or approaches or response briefs or response program*)).ti,ab. 105

21 (Timeliness adj6 (assessments or reports or qualitative research or research or studies or evaluations or appraisals or approaches or response briefs or response program*)).ti,ab. 350
22 (Timely adj (assessments or reports or qualitative research or research or studies or evaluations or appraisals or approaches or response briefs or response program*)).ti,ab. 266
23 ((Fast or faster) adj (appraisal or approach or study or assessment or report)).ti,ab. 923
24 ((Limit* or short* or tight or efficient or pragmatic) adj2 (deadline* or time or timescale or timeline) adj2 (appraisal or approach or study or assessment or report)).ti,ab. 1149
25 ((Speed or speedy) adj (appraisal or approach or study or assessment or report)).ti,ab. 196
26 (Abbreviated adj (appraisal or approach or study or assessment or report)).ti,ab. 64
27 (Accelerated adj (appraisal or approach or study or assessment or report)).ti,ab. 168
28 (Constrained by time and (appraisal or approach or study or assessment or report)).ti,ab. 33
29 (Expedited adj (appraisal or approach or study or assessment or report)).ti,ab. 61
30 (Express adj (appraisal or approach or study or assessment or report)).ti,ab. 113
31 (Immediate adj (appraisal or approach or study or assessment or report)).ti,ab. 649
32 ((Quick adj2 dirty) and (appraisal or approach or study or assessment or report)).ti,ab. 30
33 (Quick* adj (appraisal or approach or study or assessment or report)).ti,ab. 799
34 (Rapid adj (appraisal or approach or study or assessment or report)).ti,ab. 6636
35 (Stream-line* adj (appraisal or approach or study or assessment or report)).ti,ab. 5
36 (Streamline* adj (appraisal or approach or study or assessment or report)).ti,ab. 473
37 (Timeliness adj6 (appraisal or approach or study or assessment or report)).ti,ab. 556
38 (Timely adj (appraisal or approach or study or assessment or report)).ti,ab. 805
39 Participatory rural appraisal.ti,ab. 147
40 Rapid rural appraisal.ti,ab. 33
41 1 or 2 or 3 or 4 or 5 or 6 or 7 or 8 or 9 or 10 or 11 or 12 or 13 or 14 or 15 or 16 or 17 or 18 or 19 or 20 or 21 or 22 or 23 or 24 or 25 or 26 or 27 or 28 or 29 or 30 or 31 or 32 or 33 or 34 or 35 or 36 or 37 or 38 or 39 or 40 19431
42 limit 41 to yr="2002 -Current" 16894

<u>HMIC Health Management Information Consortium (searched on 11 August 2022)</u>
1 Rapid evaluation.ti,ab. 13
2 Real time evaluation.ti,ab. 3
3 Quick evaluation.ti,ab. 0
4 Short evaluation.ti,ab. 4
5 Instant evaluation.ti,ab. 0
6 Rapid cycle evaluation.ti,ab. 2
7 ((Fast or faster) adj (assessment? or report* or qualitative research or research or studies or evaluation? or appraisal? or approach* or response brief? or response program*)).ti,ab. 6
8 ((Limit* or short* or tight or efficient or pragmatic) adj2 (deadline? or time or timescale or timeline) adj2 (assessment? or report? or qualitative research or research or studies or study or evaluation? or appraisal? or approach* or response brief? or response program*)).ti,ab. 15
9 ((Speed or speedy) adj (assessments or reports or qualitative research or research or studies or evaluations or appraisals or approaches or response briefs or response program*)).ti,ab. 0
10 (Abbreviated adj (assessments or reports or qualitative research or research or studies or evaluations or appraisals or approaches or response briefs or response program*)).ti,ab. 0
11 (Accelerated adj (assessments or reports or qualitative research or research or studies or evaluations or appraisals or approaches or response briefs or response program*)).ti,ab. 0
12 (Brief adj (assessments or reports or qualitative research or research or studies or evaluations or appraisals or approaches or response program*)).ti,ab. 18
13 (Constrained by time and (assessments or reports or qualitative research or research or studies or evaluations or appraisals or approaches or response briefs or response program*)).ti,ab. 1

14 (Expedited adj (assessments or reports or qualitative research or research or studies or evaluations or appraisals or approaches or response briefs or response program*)).ti,ab. 0
15 (Express adj (assessments or reports or qualitative research or research or studies or evaluations or appraisals or approaches or response briefs or response program*)).ti,ab. 0
16 (Immediate adj (qualitative research or research or studies or evaluations or appraisals or approaches or response briefs or response program*)).ti,ab. 0
17 ((Quick adj2 dirty) and (assessments or reports or qualitative research or research or studies or evaluations or appraisals or approaches or response briefs or response program*)).ti,ab. 2
18 (Quick* adj (assessments or reports or qualitative research or research or studies or evaluations or appraisals or approaches or response briefs or response program*)).ti,ab. 3
19 (Stream-line* adj (assessments or reports or qualitative research or research or studies or evaluations or appraisals or approaches or response briefs or response program*)).ti,ab. 1
20 (Streamline* adj (assessments or reports or qualitative research or research or studies or evaluations or appraisals or approaches or response briefs or response program*)).ti,ab. 6
21 (Timeliness adj6 (assessments or reports or qualitative research or research or studies or evaluations or appraisals or approaches or response briefs or response program*)).ti,ab. 18
22 (Timely adj (assessments or reports or qualitative research or research or studies or evaluations or appraisals or approaches or response briefs or response program*)).ti,ab. 7
23 ((Fast or faster) adj (appraisal or approach or study or assessment or report)).ti,ab. 1
24 ((Limit* or short* or tight or efficient or pragmatic) adj2 (deadline* or time or timescale or timeline) adj2 (appraisal or approach or study or assessment or report)).ti,ab. 8
25 ((Speed or speedy) adj (appraisal or approach or study or assessment or report)).ti,ab. 4
26 (Abbreviated adj (appraisal or approach or study or assessment or report)).ti,ab. 2
27 (Accelerated adj (appraisal or approach or study or assessment or report)).ti,ab. 2
28 (Constrained by time and (appraisal or approach or study or assessment or report)).ti,ab. 1
29 (Expedited adj (appraisal or approach or study or assessment or report)).ti,ab. 0
30 (Express adj (appraisal or approach or study or assessment or report)).ti,ab. 0
31 (Immediate adj (appraisal or approach or study or assessment or report)).ti,ab. 12
32 ((Quick adj2 dirty) and (appraisal or approach or study or assessment or report)).ti,ab. 1
33 (Quick* adj (appraisal or approach or study or assessment or report)).ti,ab. 2
34 (Rapid adj (appraisal or approach or study or assessment or report)).ti,ab. 99
35 (Stream-line* adj (appraisal or approach or study or assessment or report)).ti,ab. 0
36 (Streamline* adj (appraisal or approach or study or assessment or report)).ti,ab. 6
37 (Timeliness adj6 (appraisal or approach or study or assessment or report)).ti,ab. 13
38 (Timely adj (appraisal or approach or study or assessment or report)).ti,ab. 7
39 Participatory rural appraisal.ti,ab. 0
40 Rapid rural appraisal.ti,ab. 0
41 1 or 2 or 3 or 4 or 5 or 6 or 7 or 8 or 9 or 10 or 11 or 12 or 13 or 14 or 15 or 16 or 17 or 18 or 19 or 20 or 21 or 22 or 23 or 24 or 25 or 26 or 27 or 28 or 29 or 30 or 31 or 32 or 33 or 34 or 35 or 36 or 37 or 38 or 39 or 40 242
42 limit 41 to yr="2002 -Current" 135

EBSCOHost CINAHL Plus (searched on 11 August 2022)
S1 Rapid evaluation Expanders - Apply equivalent subjects Search modes - Boolean/Phrase. 1,186
S2 Real time evaluation Expanders - Apply equivalent subjects Search modes - Boolean/Phrase. 544
S3 Quick evaluation Expanders - Apply equivalent subjects Search modes - Boolean/Phrase. 145
S4 Short evaluation Expanders - Apply equivalent subjects Search modes - Boolean/Phrase. 1,527
S5 Instant evaluation Expanders - Apply equivalent subjects Search modes - Boolean/Phrase. 17
S6 Rapid cycle evaluation Expanders - Apply equivalent subjects Search modes - Boolean/Phrase. 14

S7 ((Fast or faster) adj (assessment? or report* or qualitative research or research or studies or evaluation? or appraisal? or approach* or response brief? or response program*)) Expanders - Apply equivalent subjects Search modes - Boolean/Phrase. 7
S8 ((Limit* or short* or tight or efficient or pragmatic) adj2 (deadline? or time or timescale or timeline) adj2 (assessment? or report? or qualitative research or research or studies or study or evaluation? or appraisal? or approach* or response brief? or response program*)) Expanders - Apply equivalent subjects Search modes - SmartText Searching. 230
S9 ((Speed or speedy) adj (assessments or reports or qualitative research or research or studies or evaluations or appraisals or approaches or response briefs or response program*)) Expanders - Apply equivalent subjects Search modes - Boolean/Phrase. 8
S10 (Abbreviated adj (assessments or reports or qualitative research or research or studies or evaluations or appraisals or approaches or response briefs or response program*)) Expanders - Apply equivalent subjects Search modes - SmartText Searching. 670
S11 (Accelerated adj (assessments or reports or qualitative research or research or studies or evaluations or appraisals or approaches or response briefs or response program*)) Expanders - Apply equivalent subjects Search modes - SmartText Searching. 671
S12 (Brief adj (assessments or reports or qualitative research or research or studies or evaluations or appraisals or approaches or response program*)) Expanders - Apply equivalent subjects Search modes - SmartText Searching. 673
S13 ((Constrained by time) and (assessments or reports or qualitative research or research or studies or evaluations or appraisals or approaches or response briefs or response program*)) Expanders - Apply equivalent subjects Search modes - Boolean/Phrase. 254
S14 (Expedited adj (assessments or reports or qualitative research or research or studies or evaluations or appraisals or approaches or response briefs or response program*)) Expanders - Apply equivalent subjects Search modes - SmartText Searching. 670
S15 (Express adj (assessments or reports or qualitative research or research or studies or evaluations or appraisals or approaches or response briefs or response program*)) Expanders - Apply equivalent subjects Search modes - SmartText Searching. 671
S16 (Immediate adj (qualitative research or research or studies or evaluations or appraisals or approaches or response briefs or response program*)) Expanders - Apply equivalent subjects Search modes - SmartText Searching. 668
S17 ((Quick adj2 dirty) and (assessments or reports or qualitative research or research or studies or evaluations or appraisals or approaches or response briefs or response program*)) Expanders - Apply equivalent subjects Search modes - SmartText Searching. 671
S18 (Quick* adj (assessments or reports or qualitative research or research or studies or evaluations or appraisals or approaches or response briefs or response program*)) Expanders - Apply equivalent subjects Search modes - SmartText Searching. 671
S19 (Stream-line* adj (assessments or reports or qualitative research or research or studies or evaluations or appraisals or approaches or response briefs or response program*)) Expanders - Apply equivalent subjects Search modes - SmartText Searching. 675
S20 (Streamline* adj (assessments or reports or qualitative research or research or studies or evaluations or appraisals or approaches or response briefs or response program*)) Expanders - Apply equivalent subjects Search modes - SmartText Searching. 670
S21 (Timeliness adj6 (assessments or reports or qualitative research or research or studies or evaluations or appraisals or approaches or response briefs or response program*)) Expanders - Apply equivalent subjects Search modes - SmartText Searching. 671
S22 (Timely adj (assessments or reports or qualitative research or research or studies or evaluations or appraisals or approaches or response briefs or response program*)) Expanders - Apply equivalent subjects Search modes - SmartText Searching. 672
S23 ((Fast or faster) adj (appraisal or approach or study or assessment or report)) Expanders - Apply equivalent subjects Search modes - Boolean/Phrase. 7

S24 ((Limit* or short* or tight or efficient or pragmatic) adj2 (deadline* or time or timescale or timeline) adj2 (appraisal or approach or study or assessment or report)) Expanders - Apply equivalent subjects Search modes - SmartText Searching. 210
S25 ((Speed or speedy) adj (appraisal or approach or study or assessment or report)) Expanders - Apply equivalent subjects Search modes - Boolean/Phrase. 8
S26 (Abbreviated adj (appraisal or approach or study or assessment or report)) Expanders - Apply equivalent subjects Search modes - SmartText Searching. 738
S27 (Accelerated adj (appraisal or approach or study or assessment or report)) Expanders - Apply equivalent subjects Search modes - SmartText Searching. 738
S28 (Constrained by time) and (appraisal or approach or study or assessment or report)) Expanders - Apply equivalent subjects Search modes - Boolean/Phrase. 236
S29 (Expedited adj (appraisal or approach or study or assessment or report)) Expanders - Apply equivalent subjects Search modes - SmartText Searching. 738
S30 (Express adj (appraisal or approach or study or assessment or report)) Expanders - Apply equivalent subjects Search modes - SmartText Searching. 740
S31 (Immediate adj (appraisal or approach or study or assessment or report)) Expanders - Apply equivalent subjects Search modes - SmartText Searching. 739
S32 ((Quick adj2 dirty) and (appraisal or approach or study or assessment or report)) Expanders - Apply equivalent subjects Search modes - SmartText Searching. 738
S33 (Quick* adj (appraisal or approach or study or assessment or report)) Expanders - Apply equivalent subjects Search modes - SmartText Searching. 738
S34 (Rapid adj (appraisal or approach or study or assessment or report)) Expanders - Apply equivalent subjects Search modes - SmartText Searching. 741
S35 (Stream-line* adj (appraisal or approach or study or assessment or report)) Expanders - Apply equivalent subjects Search modes - SmartText Searching. 740
S36 (Streamline* adj (appraisal or approach or study or assessment or report)) Expanders - Apply equivalent subjects Search modes - SmartText Searching. 738
S37 (Timeliness adj6 (appraisal or approach or study or assessment or report)) Expanders - Apply equivalent subjects Search modes - SmartText Searching. 738
S38 (Timely adj (appraisal or approach or study or assessment or report)) Expanders - Apply equivalent subjects Search modes - SmartText Searching. 738
S39 (Participatory rural appraisal) Expanders - Apply equivalent subjects Search modes - Boolean/Phrase. 47
S40 Rapid rural appraisal Expanders - Apply equivalent subjects Search modes - Boolean/Phrase. 7
S41 S1 OR S2 OR S3 OR S4 OR S5 OR S6 OR S7 OR S8 OR S9 OR S10 OR S11 OR S12 OR S13 OR S14 OR S15 OR S16 OR S17 OR S18 OR S19 OR S20 OR S21 OR S22 OR S23 OR S24 OR S25 OR S26 OR S27 OR S28 OR S29 OR S30 OR S31 OR S32 OR S33 OR S34 OR S35 OR S36 OR S37 OR S38 OR S39 OR S40 Limiters - Publication Year: 2002-2023 Expanders - Apply equivalent subjects Search modes - Boolean/Phrase. 3,792

<u>Google Scholar (searched on 16 August 2022 – first 100 results)</u>
(rapid) and (evaluation or appraisal or approach or assessment)
Time limit: 2002-current

**Appendix 2 – Study characteristics and themes of included articles**

| First author surname | Year | Location | Type of study design | Thematic categories – methods that improved rigour and/or transparency | Quality assessment |
|---|---|---|---|---|---|
| Anker | 1993 | Botswana, Madagascar, Papua New Guinea, Uganda and Zambia | Rapid evaluation *Qualitative* | Data collection: *numerous methods for data collection; multi-disciplinary and trained researchers.*<br><br>Data analysis: *triangulation of findings.* | 2/5 |
| Aral | 2002 | Russia | Rapid assessment *Qualitative* | Data collection: *numerous methods for data collection.*<br><br>Data analysis: *iterative collection and analysis; triangulation of findings; group discussions and consensus.* | 3/5 |
| Yap | 2002 | China | Rapid assessment *Qualitative* | Study design: *consent; sampling.*<br><br>Data collection: *numerous methods for data collection; multi-disciplinary and trained researchers; recording and transcribing.*<br><br>Result interpretation: *member checking.* | 2/5 |
| Hossain | 2003 | Bangladesh | Rapid assessment *Quantitative* | Data collection: *multi-disciplinary and trained researchers.* | 3/5 |
| Miller | 2003 | Dominican Republic | Rapid assessment *Qualitative* | Data collection: *numerous methods for data collection; multi-disciplinary and trained researchers.*<br><br>Result interpretation: *advisory group support.* | 2/5 |
| Okuthe | 2003 | Kenya | Rapid rural appraisal *Mixed methods* | Data collection: *numerous methods for data collection.*<br><br>Data analysis: *triangulation of findings.* | 0/5 |

| | | | | | |
|---|---|---|---|---|---|
| Utarini | 2003 | Indonesia | Rapid assessment *Qualitative* | Frameworks.<br><br>Study design: *consent; sampling.*<br><br>Data collection: *numerous methods for data collection; local researchers and local language; multi-disciplinary and trained researchers; recording and transcribing.*<br><br>Data analysis: *content analysis; triangulation of findings.*<br><br>Result interpretation: *reflexive practice; member checking.* | 4/5 |
| Chin | 2004 | Bolivia | Rapid assessment *Qualitative* | Study design: *protocols; consent; sampling.*<br><br>Data collection: *local researchers and local language; multi-disciplinary and trained researchers; team meetings.*<br><br>Data analysis: *group discussions and consensus.* | 2/5 |
| Chung | 2004 | China | Rapid assessment *Quantitative* | Study design: *consent.*<br><br>Data collection: *piloting.*<br><br>Data analysis: *consistency and quality control.* | 2/5 |
| Goepp | 2004 | Bolivia | Rapid assessment *Qualitative* | Study design: *protocols; consent; sampling.*<br><br>Data collection: *numerous methods for data collection; local researchers and local language.*<br><br>Data analysis: *content analysis; triangulation of findings; group discussions and consensus.* | 1/5 |

| | | | | | |
|---|---|---|---|---|---|
| Hopkinson | 2004 | Kyrgyzstan | Rapid appraisal *Qualitative* | Study design: *sampling.*<br><br>Data analysis: *triangulation of findings.* | 2/5 |
| Logez | 2004 | Mongolia | Rapid assessment *Quantitative* | Study design: *sampling.*<br><br>Data collection: *multi-disciplinary and trained researchers.*<br><br>Data analysis: *triangulation of findings.* | 2/5 |
| Stajduhar | 2004 | Canada | Rapid evaluation *Qualitative* | Frameworks.<br><br>Study design: *consent; sampling; incentives.*<br><br>Data collection: *numerous methods for data collection; multi-disciplinary and trained researchers; team meetings; recording and transcribing.*<br><br>Data analysis: *ground theory approach and comparison; iterative collection and analysis; triangulation of findings; group discussions and consensus; consistency and quality control.* | 4/5 |
| Palinkas | 2004 | USA | Rapid assessment *Qualitative* | Study design: *protocols; consent; recruitment.*<br><br>Data collection: *numerous methods for data collection; recording and transcribing.*<br><br>Data analysis: *ground theory approach and comparison; triangulation of findings; consistency and quality control.* | 4/5 |
| Desmond | 2005 | Tanzania | Rapid assessment *Qualitative* | Study design: *consent; incentives.* | 1/5 |

| | | | | | |
|---|---|---|---|---|---|
| | | | | Data collection: *local researchers and local language; multi-disciplinary and trained researchers; team meetings; recording and transcribing.*<br><br>Data analysis: *content analysis.* | |
| Aral | 2005 | Russia | Rapid assessment *Qualitative* | Study design: *sampling.*<br><br>Data collection: *numerous methods for data collection.*<br><br>Data analysis: *iterative collection and analysis; group discussions and consensus.* | 0/5 |
| Shah | 2005 | South Africa | Rapid assessment *Qualitative* | Study design: *consent.*<br><br>Data collection: *local researchers and local language.* | 1/5 |
| Bayleyegn | 2006 | USA | Rapid assessment *Qualitative* | Data collection: *local researchers and local language; multi-disciplinary and trained researchers.* | 1/5 |
| Maalim | 2006 | Kenya | Participatory rural appraisal *Qualitative* | Study design: *consent; sampling.*<br><br>Data collection: *multi-disciplinary and trained researchers; participatory methods; recording and transcribing.* | 3/5 |
| Morojele | 2006 | South Africa | Rapid assessment *Qualitative* | Study design: *steering groups; consent; sampling.*<br><br>Data collection: *numerous methods for data collection; recording and transcribing.*<br><br>Data analysis: *group discussions and consensus.*<br><br>Result interpretation: *advisory group support.* | 4/5 |

| | | | | | |
|---|---|---|---|---|---|
| Croucher | 2006 | UK | Rapid appraisal<br>*Qualitative* | Study design: *consent; sampling.*<br><br>Data collection: *multi-disciplinary and trained researchers; recording and transcribing.*<br><br>Data analysis: *content analysis.* | 3/5 |
| van Kamp | 2006 | Netherlands | Rapid assessment<br>*Quantitative* | Study design: *consent.*<br><br>Data collection: *local researchers and local language.* | 3/5 |
| Rhodes | 2006 | USA | Rapid assessment<br>*Quantitative* | Data collection: *piloting.*<br><br>Data analysis: *consistency and quality control.* | 4/5 |
| Ahoua | 2007 | Democratic Republic of Congo | Rapid assessment<br>*Mixed methods* | Study design: *consent.* | 2/5 |
| Akello | 2007 | Uganda | Rapid appraisal<br>*Mixed methods* | Study design: *sampling.*<br><br>Data collection: *numerous methods for data collection.* | 2/5 |
| Pepall | 2007 | Indonesia | Participatory rapid appraisal<br>*Mixed methods* | Frameworks.<br><br>Study design: *consent; recruitment; incentives.*<br><br>Data collection: *piloting; numerous methods for data collection; local researchers and local language; multi-disciplinary and trained researchers; team meetings; recording and transcribing.* | 2/5 |

| | | | | | |
|---|---|---|---|---|---|
| | | | | Data analysis: *content analysis; triangulation of findings.* | |
| Shamsuddin | 2007 | Bangladesh | Participatory rural appraisal *Mixed methods* | Study design: *recruitment.*<br><br>Data collection: *numerous methods for data collection; participatory methods.* | 1/5 |
| Solomon | 2007 | USA | Rapid assessment *Qualitative* | Study design: *sampling.*<br><br>Data collection: *numerous methods for data collection; multi-disciplinary and trained researchers; team meetings; recording and transcribing.*<br><br>Data analysis: *content analysis; iterative collection and analysis; triangulation of findings.*<br><br>Result interpretation: *member checking.* | 4/5 |
| Chopra | 2008 | Botswana, Kenya, Malawi and Uganda | Rapid assessment *Mixed methods* | Study design: *protocols.*<br><br>Data collection: *piloting; numerous methods for data collection; multi-disciplinary and trained researchers.*<br><br>Data analysis: *consistency and quality control.* | 3/5 |
| Ash | 2008 | USA | Rapid assessment *Qualitative* | Study design: *sampling; recruitment; incentives.*<br><br>Data collection: *numerous methods for data collection; local researchers and local language; multi-disciplinary and trained researchers; team meetings; recording and transcribing.* | 5/5 |
| Balogh | 2008 | UK | Rapid appraisal *Qualitative* | Study design: *protocols; steering groups.*<br><br>Data collection: *participatory methods.* | 0/5 |

| | | | | | |
|---|---|---|---|---|---|
| | | | | Data analysis: *triangulation of findings.* | |
| Dasgupta | 2008 | India | Rapid appraisal *Qualitative* | Reporting guidelines.<br><br>Study design: *sampling.*<br><br>Data collection: *numerous methods for data collection; local researchers and local language; multi-disciplinary and trained researchers; recording and transcribing.*<br><br>Data analysis: *content analysis; consistency and quality control.* | 4/5 |
| Dozier | 2008 | Grenada | Rapid assessment *Qualitative* | Study design: *protocols; consent; sampling.*<br><br>Data collection: *numerous methods for data collection; multi-disciplinary and trained researchers; team meetings.*<br><br>Result interpretation: *advisory group support.* | 4/5 |
| Morin | 2008 | Peru, Zimbabwe, and Thailand | Rapid assessment *Qualitative* | Study design: *protocols; consent; sampling; recruitment; incentives.*<br><br>Data collection: *numerous methods for data collection; local researchers and local language; team meetings; recording and transcribing.*<br><br>Data analysis: *content analysis; group discussions and consensus.*<br><br>Result interpretation: *member checking.* | 2/5 |
| Needle | 2008 | South Africa | Rapid assessment | Frameworks. | 4/5 |

| | | | | | |
|---|---|---|---|---|---|
| | | | response and evaluation *Qualitative* | Study design: *sampling; incentives.*<br><br>Data collection: *numerous methods for data collection; multi-disciplinary and trained researchers; recording and transcribing.*<br><br>Data analysis: *content analysis.* | |
| Parry | 2008 | South Africa | Rapid assessment *Qualitative* | Study design: *protocols; consent; sampling; incentives.*<br><br>Data collection: *numerous methods for data collection; local researchers and local language; multi-disciplinary and trained researchers; recording and transcribing.*<br><br>Data analysis: *content analysis; triangulation of findings.* | 4/5 |
| Polidoro | 2008 | Costa Rica | Rapid rural appraisal *Qualitative* | Data collection: *numerous methods for data collection; local researchers and local language.*<br><br>Data analysis: *triangulation of findings.*<br><br>Result interpretation: *member checking.* | 1/5 |
| Waiswa | 2008 | Uganda | Rapid appraisal *Qualitative* | Study design: *consent.*<br><br>Data collection: *numerous methods for data collection; local researchers and local language; multi-disciplinary and trained researchers; recording and transcribing.*<br><br>Data analysis: *content analysis; triangulation of findings.* | 3/5 |
| Brown | 2008 | USA | Rapid assessment | Data collection: *numerous methods for data collection; local researchers and local language; multi-disciplinary and trained researchers; team meetings.* | 4/5 |

| | | | | | |
|---|---|---|---|---|---|
| | | | response and evaluation *Qualitative* | Data analysis: *triangulation of findings; group discussions and consensus.*<br><br>Result interpretation: *advisory group support.* | |
| Hawkins | 2009 | Mozambique | Peer ethnography *Qualitative* | Study design: *sampling.*<br><br>Data collection: *local researchers and local language; multi-disciplinary and trained researchers; recording and transcribing.* | 3/5 |
| Beckerleg | 2009 | Kenya | Rapid assessment *Quantitative* | Study design: *consent; recruitment; incentives.*<br><br>Data collection: *local researchers and local language.*<br><br>Result interpretation: *member checking.* | 1/5 |
| Betancourt | 2009 | Uganda | Rapid ethnographic assessment *Qualitative* | Study design: *sampling.*<br><br>Data collection: *local researchers and local language; multi-disciplinary and trained researchers.*<br><br>Data analysis: *iterative collection and analysis.* | 3/5 |
| Bjørkhaug | 2009 | Sierra Leone | Rapid rural assessment *Qualitative* | Study design: *sampling; incentives.*<br><br>Data collection: *local researchers and local language; multi-disciplinary and trained researchers.* | 2/5 |
| Chopra | 2009 | Kenya, Malawi and Zambia | Rapid assessment *Qualitative* | Study design: *sampling.*<br><br>Data analysis: *triangulation of findings.* | 2/5 |

| | | | | | |
|---|---|---|---|---|---|
| Dozier | 2009 | Dominican Republic | Rapid assessment *Qualitative* | Frameworks.<br><br>Study design: *protocols.*<br>Data collection: *numerous methods for data collection; local researchers and local language; multi-disciplinary and trained researchers; team meetings; recording and transcribing.*<br><br>Data analysis: *rapid analysis techniques; group discussions and consensus.* | 4/5 |
| Kahle | 2009 | USA | Rapid assessment *Quantitative* | Study design: *consent; sampling; incentives.*<br><br>Data collection: *multi-disciplinary and trained researchers.* | 3/5 |
| Liu | 2009 | China | Participatory rural appraisal *Qualitative* | Data collection: *piloting; local researchers and local language; recording and transcribing.* | 1/5 |
| Springgate | 2009 | USA | Rapid assessment *Qualitative* | Study design: *steering groups; consent.*<br><br>Data collection: *local researchers and local language.*<br><br>Data analysis: *ground theory approach and comparison.* | 3/5 |
| Turkson | 2009 | Ghana | Rapid appraisal *Mixed methods* | Study design: *sampling.*<br><br>Data collection: *piloting; local researchers and local language; multi-disciplinary and trained researchers.* | 1/5 |
| Vidal-Infer | 2009 | Spain | Rapid assessment *Mixed methods* | Frameworks.<br><br>Study design: *consent; sampling.* | 3/5 |

| | | | | | |
|---|---|---|---|---|---|
| | | | | Data collection: *numerous methods for data collection; multi-disciplinary and trained researchers.*<br><br>Data analysis: *ground theory approach and comparison; triangulation of findings.* | |
| Williams | 2009 | USA | Participatory rural appraisal<br>*Mixed methods* | Data collection: *multi-disciplinary and trained researchers; participatory methods.* | 1/5 |
| Mueller | 2010 | Niger | Participatory rural appraisal<br>*Qualitative* | Study design: *protocols.*<br><br>Data collection: *numerous methods for data collection; local researchers and local language; multi-disciplinary and trained researchers.*<br><br>Result interpretation: *advisory group support.* | 1/5 |
| Burgess-Allen | 2010 | UK | *Rapid analyses*<br>*Qualitative* | Data collection: *participatory methods.*<br><br>Data analysis: *mind mapping.*<br><br>Result interpretation: *member checking.*<br><br>Dissemination: *iterative evaluation and implementation.* | 4/5 |
| Hanvoravongchai | 2010 | Cambodia, Indonesia, Lao PDR, Taiwan, Thailand, and Vietnam | Rapid analyses<br>*Mixed methods* | Frameworks.<br><br>Study design: *sampling.*<br>Data collection: *local researchers and local language.* | 1/5 |

| | | | | | |
|---|---|---|---|---|---|
| | | | | Data analysis: *iterative collection and analysis; triangulation of findings.* | |
| Moodie | 2010 | USA | Rapid assessment *Mixed methods* | Study design: *protocols; sampling; recruitment.*<br><br>Data collection: *piloting; local researchers and local language; multi-disciplinary and trained researchers; recording and transcribing.*<br><br>Data analysis: *content analysis.* | 3/5 |
| Peltzer | 2010 | South Africa | Rapid assessment *Mixed methods* | Study design: *protocols; consent.*<br><br>Data collection: *numerous methods for data collection; recording and transcribing.*<br><br>Data analysis: *content analysis; ground theory approach and comparison; triangulation of findings; group discussions and consensus.* | 2/5 |
| Burks | 2010 | USA | Rapid assessment *Qualitative* | Frameworks.<br><br>Study design: *consent; sampling; incentives.*<br><br>Data collection: *numerous methods for data collection; multi-disciplinary and trained researchers; recording and transcribing.*<br><br>Data analysis: *content analysis; group discussions and consensus.*<br><br>Result interpretation: *peer-review of interpretations.* | 4/5 |

| | | | | | |
|---|---|---|---|---|---|
| Atuyambe | 2011 | Uganda | Rapid assessment<br>*Mixed methods* | Study design: *protocols; ethics; consent; sampling; recruitment.*<br><br>Data collection: *piloting; local researchers and local language; multi-disciplinary and trained researchers; team meetings; recording and transcribing.*<br><br>Data analysis: *iterative collection and analysis; triangulation of findings.* | 4/5 |
| Dos Santos | 2011 | South Africa | Rapid assessment response<br>*Qualitative* | Reporting guidelines.<br><br>Study design: *sampling.*<br><br>Data collection: *numerous methods for data collection; multi-disciplinary and trained researchers.*<br><br>Data analysis: *content analysis; group discussions and consensus.*<br><br>Result interpretation: *member checking.* | 1/5 |
| Ezard | 2011 | Kenya, Liberia, northern Uganda, Iran, Pakistan, and Thailand | Rapid assessment response<br>*Qualitative* | Study design: *consent; sampling.*<br><br>Data collection: *local researchers and local language; team meetings.*<br><br>Data analysis: *content analysis; iterative collection and analysis; triangulation of findings.* | 4/5 |
| Grant | 2011 | Uganda, Kenya and Malawi | Rapid evaluation<br>*Qualitative* | Frameworks.<br><br>Study design: *ethics; consent; sampling.* | 4/5 |

| | | | | | |
|---|---|---|---|---|---|
| | | | | Data collection: *numerous methods for data collection; local researchers and local language; multi-disciplinary and trained researchers; recording and transcribing.*<br><br>Data analysis: *content analysis; iterative collection and analysis; triangulation of findings; group discussions and consensus.* | |
| Kamineni | 2011 | India | Rapid assessment response *Qualitative* | Study design: *sampling.*<br><br>Data collection: *numerous methods for data collection; local researchers and local language; multi-disciplinary and trained researchers; team meetings; recording and transcribing.*<br><br>Data analysis: *triangulation of findings; group discussions and consensus.* | 4/5 |
| Maroyi | 2011 | Zimbabwe | Participatory rapid appraisal *Qualitative* | Study design: *consent.*<br><br>Data collection: *local researchers and local language.*<br><br>Data analysis: *content analysis.* | 1/5 |
| Myers | 2011 | St. Vincent and the Grenadines | Rapid appraisal *Qualitative* | Study design: *consent; sampling; incentives.*<br><br>Data collection: *numerous methods for data collection; multi-disciplinary and trained researchers; recording and transcribing.*<br><br>Data analysis: *iterative collection and analysis; triangulation of findings; group discussions and consensus.*<br><br>Result interpretation: *member checking.* | 4/5 |

| | | | | | |
|---|---|---|---|---|---|
| Poteat | 2011 | Senegal | Rapid appraisal<br>*Qualitative* | Study design: *sampling; recruitment.*<br><br>Data collection: *numerous methods for data collection; local researchers and local language; multi-disciplinary and trained researchers; team meetings; recording and transcribing.*<br><br>Data analysis: *content analysis.*<br><br>Result interpretation: *member checking.* | 4/5 |
| Kiawi | 2012 | Cameroon | Rapid assessment<br>*Qualitative* | Study design: *protocols; consent; sampling; incentives.*<br><br>Data collection: *numerous methods for data collection; local researchers and local language; multi-disciplinary and trained researchers; recording and transcribing.*<br><br>Data analysis: *content analysis; iterative collection and analysis; group discussions and consensus.* | 4/5 |
| Tindana | 2012 | Ghana | Rapid assessment<br>*Qualitative* | Study design: *consent; sampling.*<br><br>Data collection: *numerous methods for data collection; local researchers and local language; team meetings; recording and transcribing.*<br><br>Data analysis: *iterative collection and analysis.* | 2/5 |
| Mahmood, Mahmood, et al. | 2013 | Pakistan | Rapid appraisal<br>*Quantitative* | Data collection: *numerous methods for data collection; local researchers and local language.* | 1/5 |

| | | | | | |
|---|---|---|---|---|---|
| Green | 2013 | USA | Rapid assessment response *Qualitative* | Study design: *steering group; consent; sampling.*<br><br>Data collection: *multi-disciplinary and trained researchers; recording and transcribing.*<br><br>Data analysis: *iterative collection and analysis; consistency and quality control.* | 4/5 |
| Kumar | 2013 | Pakistan | Rapid appraisal *Quantitative* | Study design: *consent; sampling.*<br><br>Data collection: *piloting; multi-disciplinary and trained researchers.* | 1/5 |
| Mahmood, Rashid, et al. | 2013 | Pakistan | Rapid appraisal *Quantitative* | Study design: *recruitment.*<br><br>Data collection: *local researchers and local language.* | 1/5 |
| Turk | 2013 | Tonga | Rapid assessment response *Qualitative* | Data collection: *numerous methods for data collection; multi-disciplinary and trained researchers; recording and transcribing.*<br><br>Data analysis: g*round theory approach and comparison; triangulation of findings.* | 2/5 |
| Van Hout | 2013 | Republic of Ireland | Rapid assessment *Qualitative* | Study design: *consent.*<br><br>Data collection: *numerous methods for data collection; recording and transcribing.*<br><br>Data analysis: *content analysis.*<br><br>Result interpretation: *reflexive practice.* | 4/5 |

| | | | | | |
|---|---|---|---|---|---|
| Wan | 2013 | China | Rapid assessment *Quantitative* | Study design: *protocols; consent.*<br><br>Data collection: *numerous methods for data collection; multi-disciplinary and trained researchers.*<br><br>Data analysis: *consistency and quality control.* | 4/5 |
| Dwyer | 2014 | Australia | Rapid assessment *Mixed methods* | Frameworks.<br><br>Study design: *consent; recruitment; incentives.*<br><br>Data collection: *numerous methods for data collection; recording and transcribing.*<br><br>Data analysis: *content analysis; triangulation of findings.* | 1/5 |
| Joarder | 2014 | Bangladesh | Participatory rapid appraisal *Qualitative* | Study design: *consent; sampling; recruitment.*<br><br>Data collection: *numerous methods for data collection; local researchers and local language; recording and transcribing.*<br><br>Data analysis: *content analysis.* | 3/5 |
| Kyamwanga | 2014 | Uganda | Rapid assessment *Qualitative* | Study design: *sampling; incentives.*<br><br>Data collection: *numerous methods for data collection; recording and transcribing.*<br><br>Data analysis: *content analysis; triangulation of findings.* | 2/5 |
| Leonard | 2014 | South Africa | Rapid appraisal *Qualitative* | Study design: *consent.*<br><br>Data analysis: *content analysis; iterative collection and analysis.* | 3/5 |

| | | | | | |
|---|---|---|---|---|---|
| | | | | Result interpretation: *peer-review of interpretations.* | |
| Oloukoi | 2014 | Nigeria | Rapid appraisal *Mixed methods* | Study design: *sampling.*<br><br>Data collection: *numerous methods for data collection; participatory methods.*<br><br>Data analysis: *content analysis.* | 2/5 |
| Wilunda | 2014 | Uganda | Participatory rural appraisal *Qualitative* | Study design: *consent; sampling; incentives.*<br><br>Data collection: *local researchers and local language; multi-disciplinary and trained researchers; participatory methods; recording and transcribing.*<br><br>Data analysis: *content analysis; triangulation of findings.*<br><br>Result interpretation: *member checking.* | 4/5 |
| Ahmed, Mahmood, Ashraf, et al. | 2015 | Pakistan | Rapid appraisal *Mixed methods* | Data collection: *numerous methods for data collection; local researchers and local language.* | 2/5 |
| Ahmed, Mahmood, Mahmood, et al. | 2015 | Pakistan | Rapid appraisal *Mixed methods* | Data collection: *numerous methods for data collection; local researchers and local language.* | 2/5 |
| Barden-O'Fallon | 2015 | Guinea | Rapid assessment *Qualitative* | Study design: *ethics; consent.*<br><br>Data collection: *piloting; multi-disciplinary and trained researchers* | 4/5 |

| | | | | | |
|---|---|---|---|---|---|
| | | | | Data analysis: *consistency and quality control.* | |
| Kodish, Aburto, Dibari, et al. | 2015 | Mozambique | Rapid assessment *Qualitative* | Study design: *protocols; consent; sampling.*<br><br>Data collection: *piloting; numerous methods for data collection; local researchers and local language; multi-disciplinary and trained researchers; recording and transcribing.*<br><br>Data analysis: *iterative collection and analysis; triangulation of findings.*<br><br>Result interpretation: *member checking.* | 5/5 |
| Kodish, Aburto, Hambayi, et al. | 2015 | Malawi | Rapid assessment *Qualitative* | Study design: *protocols; consent; sampling; recruitment.*<br><br>Data collection: *numerous methods for data collection; local researchers and local language.*<br><br>Result interpretation: *member checking.* | 4/5 |
| Sambo | 2015 | Ethiopia | Participatory evaluation *Mixed methods* | Study design: *consent.*<br><br>Data collection: *numerous methods for data collection; local researchers and local language; participatory methods.*<br><br>Data analysis: *triangulation of findings.* | 1/5 |
| Seay | 2015 | USA | Rapid assessment *Quantitative* | Study design: *protocols; steering group.*<br><br>Data collection: *local researchers and local language; multi-disciplinary and trained researchers.* | 2/5 |

| | | | | | |
|---|---|---|---|---|---|
| Mital | 2016 | Kenya | Rapid assessment *Qualitative* | Study design: *consent; sampling; incentives.*<br><br>Data collection: *numerous methods for data collection; local researchers and local language; multi-disciplinary and trained researchers; recording and transcribing.*<br><br>Data analysis: *content analysis; triangulation of findings; consistency and quality control.* | 4/5 |
| Ash | 2016 | USA | Rapid assessment *Qualitative* | Study design: *protocols; recruitment;*<br><br>Data collection: *numerous methods for data collection; multi-disciplinary and trained researchers; team meetings.*<br><br>Data analysis: *ground theory approach and comparison; iterative collection and analysis; group discussions and consensus.*<br><br>Result interpretation: *team consensus; member checking.* | 5/5 |
| Aziz | 2016 | Pakistan | Rapid appraisal *Mixed methods* | Data collection: *numerous methods for data collection; local researchers and local language.* | 1/5 |
| Livorsi | 2016 | USA | Rapid assessment *Qualitative* | Frameworks.<br><br>Study design: *sampling.*<br><br>Data collection: *recording and transcribing.*<br><br>Data analysis: *content analysis.* | 4/5 |

| | | | | | |
|---|---|---|---|---|---|
| Theiss-Nyland | 2016 | Kenya, Malawi, Mali and Rwanda | Rapid assessment<br>*Qualitative* | Study design: *protocols; consent; sampling.*<br><br>Data collection: *numerous methods for data collection; local researchers and local language; multi-disciplinary and trained researchers; recording and transcribing.*<br><br>Data analysis: *triangulation of findings.* | 2/5 |
| Weiss | 2016 | Zambia | Rapid assessment<br>*Qualitative* | Study design: *consent.*<br><br>Data collection: *piloting; numerous methods for data collection; local researchers and local language; multi-disciplinary and trained researchers.*<br><br>Data analysis: g*round theory approach and comparison; iterative collection and analysis.* | 4/5 |
| Altaras | 2017 | Uganda | Rapid appraisal<br>*Qualitative* | Frameworks*.*<br><br>Study design: *protocols; sampling.*<br><br>Data collection: *piloting; local researchers and local language; multi-disciplinary and trained researchers; participatory methods; recording and transcribing.*<br><br>Data analysis: *content analysis.*<br><br>Result interpretation: *member checking.* | 4/5 |
| Belackova | 2017 | Czech Republic | Rapid assessment response | Frameworks*.* | 2/5 |

| | | | | | |
|---|---|---|---|---|---|
| | | | *Mixed methods* | | |
| Belford | 2017 | UK | Evaluation *Qualitative* | Data analysis: *iterative collection and analysis; group discussions and consensus.*<br><br>Result interpretation: *team consensus; member checking.* | 2/5 |
| Doherty | 2017 | Uganda | Rapid appraisal *Qualitative* | Frameworks.<br><br>Study design: *protocols; consent.*<br><br>Data collection: *numerous methods for data collection; local researchers and local language; recording and transcribing.*<br><br>Data analysis: *triangulation of findings.* | 5/5 |
| Kraaij-Dirkzwager | 2017 | Netherlands | Rapid assessment *Qualitative* | Study design: *consent; sampling.*<br><br>Data collection: *recording and transcribing.*<br><br>Data analysis: *ground theory approach and comparison; group discussions and consensus.* | 4/5 |
| Negandhi | 2017 | India | Rapid assessment *Qualitative* | Study design: *protocols; sampling.*<br><br>Data collection: *local researchers and local language; multi-disciplinary and trained researchers; recording and transcribing.*<br><br>Data analysis: *content analysis.* | 4/5 |
| Turk | 2017 | Vietnam | Rapid assessment *Qualitative* | Study design: *sampling.* | 4/5 |

| | | | | | |
|---|---|---|---|---|---|
| | | | | Data collection: *numerous methods for data collection; multi-disciplinary and trained researchers; recording and transcribing.*<br><br>Data analysis: *ground theory approach and comparison.* | |
| Van Meer | 2017 | Canada | Evaluation<br>*Qualitative* | Data collection: *recording and transcribing.*<br><br>Data analysis: *content analysis.*<br><br>Result interpretation: *member checking.* | 1/5 |
| Wright | 2017 | USA | Rapid assessment<br>*Qualitative* | Data collection: *multi-disciplinary and trained researchers; recording and transcribing.*<br><br>Data analysis: *ground theory approach and comparison.* | 4/5 |
| Harapan | 2018 | Indonesia | Rapid assessment<br>*Quantitative* | Study design: *protocols; consent; recruitment; incentives.*<br><br>Data collection: *piloting; technology.* | 3/5 |
| Guise | 2018 | UK | Rapid assessment<br>*Qualitative* | Study design: *consent; sampling; incentives.*<br><br>Data collection: *numerous methods for data collection.*<br><br>Data analysis: *content analysis; triangulation of findings.* | 4/5 |
| Murphy | 2018 | Georgia | Rapid appraisal<br>*Qualitative* | Study design: *consent; sampling.*<br><br>Data collection: *numerous methods for data collection; local researchers and local language; multi-disciplinary and trained researchers; recording and transcribing.*<br><br>Data analysis: *content analysis; triangulation of findings.* | 2/5 |

| | | | | | |
|---|---|---|---|---|---|
| Neal | 2018 | Haiti | Participatory rural appraisal *Qualitative* | Study design: *incentives.*<br><br>Data collection: *local researchers and local language; multi-disciplinary and trained researchers; participatory methods.*<br><br>Result interpretation: *team consensus; member checking.* | 2/5 |
| Nemser | 2018 | Malawi | Rapid appraisal *Qualitative* | Study design: *consent; sampling.*<br><br>Data collection: *local researchers and local language; recording and transcribing.*<br><br>Data analysis: *content analysis; group discussions and consensus.* | 2/5 |
| Nsibande | 2018 | Ethiopia | Rapid appraisal *Qualitative* | Study design: *protocols; consent.*<br><br>Data collection: *local researchers and local language; multi-disciplinary and trained researchers; recording and transcribing.*<br><br>Data analysis: *content analysis; group discussions and consensus.* | 4/5 |
| Seidel | 2018 | Kenya | Rapid rural appraisal *Qualitative* | Study design: *sampling.*<br><br>Data collection: *local researchers and local language; multi-disciplinary and trained researchers; recording and transcribing.*<br><br>Data analysis: *content analysis.*<br><br>Result interpretation: *member checking.* | 4/5 |

| | | | | | |
|---|---|---|---|---|---|
| Barreras | 2019 | USA | Rapid assessment *Qualitative* | Study design: *consent; recruitment; incentives.*<br><br>Data collection: *multi-disciplinary and trained researchers.*<br><br>Data analysis: *rapid analysis techniques.* | 4/5 |
| Brittain | 2019 | USA | Rapid assessment *Qualitative* | Study design: *protocols; ethics; consent; recruitment; incentives.*<br><br>Data collection: *piloting; multi-disciplinary and trained researchers; recording and transcribing.*<br><br>Data analysis: *iterative collection and analysis; group discussions and consensus; consistency and quality control.* | 3/5 |
| Foreman-Mackey | 2019 | Canada | Rapid evaluation *Qualitative* | Study design: *consent; sampling; incentives.*<br><br>Data collection: *recording and transcribing.*<br><br>Data analysis: *content analysis; iterative collection and analysis.* | 4/5 |
| Hipgrave | 2019 | Bangladesh, Indonesia, Nepal and the Philippines | Rapid assessment *Qualitative* | Data collection: *numerous methods for data collection; recording and transcribing.*<br><br>Data analysis: *triangulation of findings.* | 1/5 |
| Kibe | 2019 | Kenya | Participatory rural appraisal *Qualitative* | Study design: *protocols; consent; sampling.*<br><br>Data collection: *numerous methods for data collection; local researchers and local language; multi-disciplinary and trained researchers; team meetings; recording and transcribing.* | 4/5 |

| | | | | | |
|---|---|---|---|---|---|
| | | | | Data analysis: *content analysis; ground theory approach and comparison; triangulation of findings; consistency and quality control.* | |
| Loko | 2019 | Republic of Benin | Participatory rural appraisal *Quantitative* | Study design: *consent; sampling.*<br><br>Data collection: *numerous methods for data collection; local researchers and local language.*<br><br>Data analysis: *triangulation of findings.* | 2/5 |
| Mullane | 2019 | USA | Rapid assessment *Qualitative* | Frameworks.<br><br>Study design: *consent; incentives.*<br><br>Data collection: *numerous methods for data collection; recording and transcribing.*<br><br>Data analysis: *content analysis.* | 1/5 |
| Zhou | 2019 | China | Rapid appraisal *Mixed methods* | Study design: *consent.*<br><br>Data collection: *numerous methods for data collection; recording and transcribing.*<br><br>Data analysis: *content analysis; triangulation of findings; consistency and quality control.* | 3/5 |
| Attal | 2020 | Israel | Rapid assessment *Quantitative* | Study design: *sampling; recruitment.*<br><br>Data collection: *technology.* | 2/5 |

| | | | | | |
|---|---|---|---|---|---|
| Dainty | 2020 | Canada | Rapid ethnographic evaluation *Qualitative* | Study design: *consent; sampling.*<br><br>Data collection: *numerous methods for data collection; multi-disciplinary and trained researchers; recording and transcribing.*<br><br>Data analysis: *content analysis; iterative collection and analysis; group discussions and consensus; audit trail.* | 4/5 |
| Gao | 2020 | China | Rapid assessment *Quantitative* | Study design: *consent.*<br><br>Data collection: *technology.* | 3/5 |
| Holdsworth | 2020 | USA | Rapid assessment *Qualitative* | Frameworks.<br><br>Data collection: *numerous methods for data collection; multi-disciplinary and trained researchers; team meetings; participatory methods; recording and transcribing.*<br><br>Data analysis: *iterative collection and analysis; triangulation of findings; group discussions and consensus.*<br><br>Result interpretation: *team consensus; member checking.* | 5/5 |
| Jack | 2020 | UK | Evaluation *Qualitative* | Study design: *consent; sampling.*<br><br>Data collection: *multi-disciplinary and trained researchers; participatory methods.*<br><br>Data analysis: *content analysis; group discussions and consensus.*<br><br>Result interpretation: *member checking.* | 4/5 |

| | | | | | |
|---|---|---|---|---|---|
| Moloney | 2020 | USA | Rapid assessment procedure informed clinical ethnography *Qualitative* | Frameworks.<br><br>Data collection: *numerous methods for data collection; multi-disciplinary and trained researchers; recording and transcribing.*<br><br>Data analysis: *content analysis.* | 1/5 |
| Naylor | 2020 | UK | Rapid assessment *Mixed methods* | Study design: *recruitment; incentives.*<br><br>Data collection: *piloting; technology.*<br><br>Data analysis: *content analysis.* | 2/5 |
| Rahman | 2020 | Bangladesh | Rapid assessment *Quantitative* | Study design: *protocols; consent; sampling.*<br><br>Data collection: *local researchers and local language; multi-disciplinary and trained researchers; team meetings.* | 2/5 |
| Sharma | 2020 | USA | Rapid assessment *Mixed methods* | Study design: *consent.*<br><br>Data collection: *local researchers and local language; technology.*<br><br>Data analysis: *content analysis; consistency and quality control.* | 3/5 |
| Sy | 2020 | Micronesia | Rapid assessment *Qualitative* | Study design: *sampling.*<br><br>Data collection: *piloting; numerous methods for data collection; team meetings; recording and transcribing.*<br><br>Data analysis: *content analysis; triangulation of findings.* | 4/5 |

| | | | | | |
|---|---|---|---|---|---|
| Vindrola-Padros | 2020 | UK | Rapid appraisal *Qualitative* | Study design: *protocols; sampling.*<br><br>Data collection: *numerous methods for data collection; multi-disciplinary and trained researchers; recording and transcribing.*<br><br>Data analysis: *rapid analysis techniques.*<br><br>Dissemination: *iterative evaluation and implementation.* | 1/5 |
| Gaiha | 2021 | India | Rapid evaluation *Mixed methods* | Study design: *consent.*<br><br>Data collection: *numerous methods for data collection; local researchers and local language; recording and transcribing.* | 2/5 |
| Robles | 2021 | USA | Rapid assessment *Qualitative* | Frameworks.<br><br>Study design: *sampling; recruitment.*<br><br>Data collection: *technology; recording and transcribing.*<br><br>Data analysis: *content analysis; group discussions and consensus.* | 4/5 |
| Fadholah | 2021 | Indonesia | Rapid assessment *Qualitative* | Study design: *sampling.*<br><br>Data collection: *recording and transcribing.*<br><br>Data analysis: *content analysis.*<br><br>Result interpretation: *reflexive practice.* | 1/5 |
| Butler | 2021 | 11 Indo-Pacific countries (the | Rapid assessment | Frameworks. | 1/5 |

| | | | | | |
|---|---|---|---|---|---|
| | | Philippines, Indonesia, Timor-Leste, Papua New Guinea (PNG) and seven Pacific Island Countries) | *Mixed methods* | Study design: *consent; sampling; recruitment.*<br><br>Data collection: *multi-disciplinary and trained researchers; team meetings; recording and transcribing.*<br><br>Data analysis: *triangulation of findings.*<br><br>Result interpretation: *peer-review of interpretations.* | |
| Abir | 2021 | USA | Rapid assessment *Qualitative* | Data analysis: *rapid analysis techniques; group discussions and consensus.* | 4/5 |
| Al Awaidy | 2021 | Gulf Cooperation Council Member States - Bahrain, Kuwait, Qatar, Oman and Saudia Arabia | Rapid appraisal *Quantitative* | Data analysis: *triangulation of findings.* | 1/5 |
| Albert | 2021 | USA | Rapid assessment *Qualitative* | Study design: *protocols; incentives.*<br><br>Data collection: *multi-disciplinary and trained researchers; technology; recording and transcribing.*<br><br>Data analysis: *iterative collection and analysis; group discussions and consensus; consistency and quality control.* | 5/5 |
| Carter-Edwards | 2021 | USA | Rapid assessment *Qualitative* | Study design: *consent; incentives.*<br><br>Data collection: *recording and transcribing.*<br><br>Data analysis: *rapid analysis techniques; group discussions and consensus; consistency and quality control.* | 5/5 |

| | | | | | |
|---|---|---|---|---|---|
| Cohn | 2021 | USA | Rapid evaluation<br>*Qualitative* | Frameworks.<br><br>Data analysis: *iterative collection and analysis; group discussions and consensus.*<br><br>Dissemination: *iterative evaluation and implementation.* | 5/5 |
| Douglass | 2021 | India | Rapid assessment<br>*Mixed methods* | Study design: *consent; sampling.*<br><br>Data collection: *piloting; multi-disciplinary and trained researchers; recording and transcribing.*<br><br>Data analysis: *rapid analysis techniques; consistency and quality control.* | 1/5 |
| Jumbe | 2021 | UK | Rapid evaluation<br>*Qualitative* | Study design: *consent; sampling; incentives.*<br><br>Data collection: *multi-disciplinary and trained researchers; technology; recording and transcribing.*<br><br>Data analysis: *content analysis; iterative collection and analysis; triangulation of findings; group discussions and consensus.*<br><br>Result interpretation: *reflexive practice.* | 4/5 |
| Kalil | 2021 | Ethiopia | Evaluation<br>*Mixed methods* | Study design: *sampling.*<br><br>Data collection: *multi-disciplinary and trained researchers.*<br><br>Data analysis: *content analysis; triangulation of findings.* | 1/5 |

| | | | | | |
|---|---|---|---|---|---|
| Laisser | 2021 | Malawi | Rapid evaluation *Qualitative* | Study design: *ethics; consent; sampling.*<br><br>Data collection: *multi-disciplinary and trained researchers; recording and transcribing.*<br><br>Data analysis: *content analysis; audit trail; consistency and quality control.* | 4/5 |
| Lynch | 2021 | Nigeria | Participatory needs assessment *Mixed methods* | Study design: *protocols; consent.*<br><br>Data collection: *numerous methods for data collection; local researchers and local language; recording and transcribing.*<br><br>Data analysis: *content analysis; triangulation of findings; group discussions and consensus.* | 2/5 |
| Mitchinson | 2021 | UK | Rapid appraisal *Qualitative* | Study design: *consent; sampling; recruitment.*<br><br>Data collection: *numerous methods for data collection; multi-disciplinary and trained researchers; recording and transcribing.*<br><br>Data analysis: *content analysis; iterative collection and analysis; triangulation of findings; group discussions and consensus.*<br><br>Result interpretation: *member checking.* | 4/5 |
| Palinkas | 2021 | USA | Rapid assessment *Qualitative* | Frameworks.<br><br>Study design: *consent; sampling; recruitment.*<br><br>Data collection: *technology; recording and transcribing.* | 4/5 |

| | | | | | |
|---|---|---|---|---|---|
| | | | | Data analysis: *content analysis; consistency and quality control.* | |
| Quach | 2021 | USA | Rapid assessment *Quantitative* | Study design: *ethics; consent; recruitment; incentives.*<br><br>Data collection: *piloting; local researchers and local language; technology.* | 3/5 |
| Shimkhada | 2021 | USA | Rapid assessment *Qualitative* | Study design: *ethics.*<br><br>Data collection: *technology.* | 4/5 |
| Sithole | 2021 | Zimbabwe | Rapid assessment *Quantitative* | Study design: *sampling.*<br><br>Data collection: *numerous methods for data collection; multi-disciplinary and trained researchers.*<br><br>Data analysis: *triangulation of findings.* | 3/5 |
| Tort-Nasarre | 2021 | Spain | Rapid appraisal *Qualitative* | Frameworks.<br><br>Reporting guidelines.<br><br>Study design: *consent; sampling; recruitment.*<br><br>Data collection: *technology; recording and transcribing.*<br><br>Data analysis: *content analysis; iterative collection and analysis.*<br><br>Result interpretation: *reflective practice.* | 4/5 |
| Tran | 2021 | Vietnam | Rapid assessment *Quantitative* | Study design: *sampling.*<br><br>Data collection: *technology.* | 3/5 |

| | | | | | |
|---|---|---|---|---|---|
| van der Merwe | 2021 | South Africa | Rapid evaluation<br>*Qualitative* | Study design: *protocols; sampling.*<br><br>Data collection: *numerous methods for data collection; recording and transcribing.*<br><br>Data analysis: *content analysis.*<br><br>Result interpretation: *member checking.* | 3/5 |
| von Thiele Schwarz | 2021 | Sweden | Rapid evaluation<br>*Mixed methods* | Study design: *consent; recruitment.*<br><br>Data collection: *numerous methods for data collection; participatory methods; recording and transcribing.*<br><br>Data analysis: *content analysis.* | 4/5 |
| Palinkas | 2022 | USA | Rapid ethnographic assessment<br>*Qualitative* | Frameworks.<br><br>Study design: *protocols; consent.*<br><br>Data collection: *numerous methods for data collection; multi-disciplinary and trained researchers; recording and transcribing.*<br><br>Data analysis: *content analysis.*<br><br>Result interpretation: *member checking.* | 4/5 |
| Anastasaki | 2022 | Greece | Rapid assessment<br>*Qualitative* | Frameworks.<br><br>Reporting guidelines. | 4/5 |

| | | | | | |
|---|---|---|---|---|---|
| | | | | Study design: *protocols; sampling; recruitment.*<br><br>Data collection: *numerous methods for data collection; local researchers and local language; multi-disciplinary and trained researchers; team meetings.*<br><br>Data analysis: *iterative collection and analysis; triangulation of findings; group discussions and consensus.* | |
| Austin | 2022 | USA | Rapid assessment *Qualitative* | Frameworks.<br><br>Data collection: *multi-disciplinary and trained researchers; technology.*<br><br>Data analysis: *rapid analysis techniques; triangulation of findings; group discussions and consensus.* | 4/5 |
| Balde | 2022 | Guinea | Rapid assessment *Mixed methods* | Study design: *protocols; consent; sampling.*<br><br>Data collection: *numerous methods for data collection; local researchers and local language; multi-disciplinary and trained researchers; recording and transcribing.* | 2/5 |
| Chilanga | 2022 | Malawi | Rapid appraisal *Qualitative* | Frameworks.<br><br>Study design: *protocols; consent; sampling; recruitment.*<br><br>Data collection: *piloting; numerous methods for data collection; local researchers and local language; multi-disciplinary and trained researchers; technology.*<br><br>Data analysis: *iterative collection and analysis; consistency and quality control.* | 5/5 |

| | | | | | |
|---|---|---|---|---|---|
| | | | | Result interpretation: *member checking.* | |
| Chung | 2022 | Canada | Rapid evaluation<br>*Qualitative* | Frameworks.<br><br>Study design: *protocols; consent; recruitment.*<br><br>Data analysis: *triangulation of findings; group discussions and consensus; consistency and quality control.*<br><br>Result interpretation: *member checking.* | 5/5 |
| Delobelle | 2022 | South Africa | Rapid appraisal<br>*Mixed methods* | Study design: *consent; sampling.*<br><br>Data collection: *local researchers and local language; multi-disciplinary and trained researchers; technology.*<br><br>Data analysis: *content analysis; iterative collection and analysis; triangulation of findings; group discussions and consensus.* | 2/5 |
| Gawaya | 2022 | Australia | Rapid evaluation<br>*Mixed methods* | Study design: *steering group; consent; sampling.*<br><br>Data collection: *numerous methods for data collection; local researchers and local language; technology; recording and transcribing.*<br><br>Data analysis: *Rigorous and Accelerated Data Reduction (RADaR) technique; iterative collection and analysis; triangulation of findings; group discussions and consensus.*<br><br>Result interpretation: *advisory group support.* | 1/5 |

| | | | | | |
|---|---|---|---|---|---|
| Hammond | 2022 | UK | Rapid assessment *Mixed methods* | Study design: *consent; sampling; recruitment.*<br><br>Data collection: *piloting; technology.* | 4/5 |
| Kumar | 2022 | Kenya | Rapid appraisal *Qualitative* | Study design: *consent.*<br><br>Data collection: *piloting; participatory methods; technology; recording and transcribing.* | 2/5 |
| Lim | 2022 | Malaysia | Rapid assessment *Mixed methods* | Study design: *consent; sampling; recruitment.*<br><br>Data collection: *piloting; local researchers and local language; recording and transcribing.*<br><br>Data analysis: *content analysis; triangulation of findings.* | 2/5 |
| Luz | 2022 | Brazil | Rapid evaluation *Mixed methods* | Study design: *protocols; sampling.*<br><br>Data collection: *piloting; numerous methods for data collection; multi-disciplinary and trained researchers.*<br><br>Data analysis: *content analysis; triangulation of findings.* | 3/5 |

| | | | | | |
|---|---|---|---|---|---|
| O'Meara | 2022 | 119 countries | Rapid assessment *Mixed methods* | Frameworks.<br><br>Study design: *ethics; consent; sampling; recruitment.*<br><br>Data collection: *piloting; technology.*<br><br>Data analysis: *content analysis; mind mapping; triangulation of findings; group discussions and consensus.* | 1/5 |
| Romeu-Labayen | 2022 | Spain | Rapid appraisal *Qualitative* | Frameworks.<br><br>Reporting guidelines.<br><br>Study design: *consent; sampling.*<br><br>Data collection: *multi-disciplinary and trained researchers; technology; recording and transcribing.*<br><br>Data analysis: *content analysis; group discussions and consensus.*<br><br>Result interpretation: *reflexive practice.* | 4/5 |
| Walton | 2022 | UK | Rapid assessment *Mixed methods* | Study design: *protocols; consent.*<br><br>Data collection: *piloting; numerous methods for data collection; local researchers and local language; multi-disciplinary and trained researchers; technology.* | 2/5 |

| | | | | | |
|---|---|---|---|---|---|
| | | | | Data analysis: *content analysis; rapid analysis techniques; triangulation of findings.* | |
| Forber-Pratt | 2022 | USA | Rapid assessment *Mixed methods* | Study design: *recruitment.*<br><br>Data collection: *technology.*<br><br>Data analysis: *rapid analysis techniques; consistency and quality control.* | 4/5 |
| Rinehart | 2023 (Epub 2022) | USA | Rapid assessment *Qualitative* | Reporting guidelines.<br><br>Study design: *protocols; consent; sampling; recruitment; incentives.*<br><br>Data collection: *multi-disciplinary and trained researchers; recording and transcribing.*<br><br>Data analysis: *rapid analysis techniques; group discussions and consensus.*<br><br>Result interpretation: *advisory group support.* | 4/5 |
| Corcorran | 2023 (Epub 2022) | USA | Rapid assessment *Qualitative* | Study design: *protocols.*<br><br>Data analysis: *rapid analysis techniques; iterative collection and analysis; group discussions and consensus.*<br><br>Result interpretation: *member checking.* | 4/5 |

**Appendix 3 – References of included items**


Abir, M., Forman, J., Taymour, R. K., Brent, C., Nallamothu, B. K., Scott, J., & Wahl, K. (2021). Optimizing Routine and Disaster Prehospital Care Through Improved Emergency Medical Services Oversight. *Disaster Medicine and Public Health Preparedness*, *15*(5), 595–607. https://doi.org/10.1017/dmp.2020.71

Ahmed, N., Mahmood, A., Ashraf, A., Bano, A., Tahir, S. S., & Mahmood, A. (2015). Ethnopharmacological relevance of indigenous medicinal plants from district Bahawalnagar, Punjab, Pakistan. *Journal of Ethnopharmacology*, *175*, 109–123. https://doi.org/10.1016/j.jep.2015.08.011

Ahmed, N., Mahmood, A., Mahmood, A., Sadeghi, Z., & Farman, M. (2015). Ethnopharmacological importance of medicinal flora from the district of Vehari, Punjab province, Pakistan. *Journal of Ethnopharmacology*, *168*, 66–78. https://doi.org/10.1016/j.jep.2015.02.048

Ahoua, L., Tamrat, A., Duroch, F., Grais, R. F., & Brown, V. (2006). High mortality in an internally displaced population in Ituri, Democratic Republic of Congo, 2005: results of a rapid assessment under difficult conditions. *Global Public Health*, *1*(3), 195–204. https://doi.org/10.1080/17441690600681869

Akello, G., Reis, R., Ovuga, E., Rwabukwali, C. B., Kabonesa, C., & Richters, A. (2007). Primary school children's perspectives on common diseases and medicines used: implications for school healthcare programmes and priority setting in Uganda. *African Health Sciences*, *7*(2), 73–79. https://doi.org/10.5555/afhs.2007.7.2.73

Al Awaidy, S. T., Khamis, F., Al Attar, F., Razzaq, N. A., Al Dabal, L., Al Enani, M., Alfouzan, W., Al Maslamani, M., Al Romaihi, H., Al Salman, J., Altawalah, H., Langrial, S. U., Al Ariqi, L., & Mohamed, O. (2021). COVID-19 in the Gulf Cooperation Council Member States: An Evidence of Effective Response. *Oman Medical Journal*, *36*(5), e300. https://doi.org/10.5001/omj.2021.115

Albert, S. L., Paul, M. M., Nguyen, A. M., Shelley, D. R., & Berry, C. A. (2021). A qualitative study of high-performing primary care practices during the COVID-19 pandemic. *BMC Family Practice*, *22*(1), 237. https://doi.org/10.1186/s12875-021-01589-4

Altaras, R., Montague, M., Graham, K., Strachan, C. E., Senyonjo, L., King, R., Counihan, H., Mubiru, D., Källander, K., Meek, S., & Tibenderana, J. (2017). Integrated community case management in a peri-urban setting: a qualitative evaluation in Wakiso District, Uganda. *BMC Health Services Research*, *17*(1), 785. https://doi.org/10.1186/s12913-017-2723-0

Anastasaki, M., van Bree, E. M., Brakema, E. A., Tsiligianni, I., Sifaki-Pistolla, D., Chatzea, V. E., Crone, M. C., Karelis, A., van der Kleij, R. M. J. J., Poot, C. C., Reis, R., Chavannes, N. H., & Lionis, C. (2022). Beliefs, Perceptions, and Behaviors Regarding Chronic Respiratory Diseases of Roma in Crete, Greece: A Qualitative FRESH AIR Study. *Frontiers in Public Health*, *10*, 812700. https://doi.org/10.3389/fpubh.2022.812700

Anker, M., Guidotti, R. J., Orzeszyna, S., Sapirie, S. A., & Thuriaux, M. C. (1993). Rapid evaluation methods (REM) of health services performance: methodological observations. *Bulletin of the World Health Organization*, *71*(1), 15–21.

Aral, S. O., & St. Lawrence, J. S. (2002). The Ecology of Sex Work and Drug Use in Saratov Oblast, Russia. *Sexually Transmitted Diseases*, *29*(12), 798–805. https://doi.org/10.1097/00007435-200212000-00011

Aral, S. O., St Lawrence, J. S., Dyatlov, R., & Kozlov, A. (2005). Commercial sex work, drug use, and sexually transmitted infections in St. Petersburg, Russia. *Social Science & Medicine (1982)*, *60*(10), 2181–2190. https://doi.org/10.1016/j.socscimed.2004.10.009

Ash, J. S., Chase, D., Wiesen, J. F., Murphy, E. V, & Marovich, S. (2016). Studying Readiness for Clinical Decision Support for Worker Health Using the Rapid Assessment Process and Mixed Methods Interviews. *AMIA ... Annual Symposium Proceedings. AMIA Symposium*, *2016*, 285–294.

Ash, J. S., Sittig, D. F., McMullen, C. K., Guappone, K., Dykstra, R., & Carpenter, J. (2008). A rapid assessment process for clinical informatics interventions. *AMIA ... Annual Symposium Proceedings. AMIA Symposium*, *2008*, 26–30.

Attal, J. H., Lurie, I., & Neumark, Y. (2020). A rapid assessment of migrant careworkers' psychosocial status during Israel's COVID-19 lockdown. *Israel Journal of Health Policy Research*, *9*(1), 61. https://doi.org/10.1186/s13584-020-00422-0

Atuyambe, L. M., Ediau, M., Orach, C. G., Musenero, M., & Bazeyo, W. (2011). Land slide disaster in eastern Uganda: rapid assessment of water, sanitation and hygiene situation in Bulucheke camp, Bududa district. *Environmental Health : A Global Access Science Source*, *10*, 38. https://doi.org/10.1186/1476-069X-10-38

Austin, E. J., Tsui, J. I., Barry, M. P., Tung, E., Glick, S. N., Ninburg, M., & Williams, E. C. (2022). Health care-seeking experiences for people who inject drugs with hepatitis C: Qualitative explorations of stigma. *Journal of Substance Abuse Treatment*, *137*, 108684. https://doi.org/10.1016/j.jsat.2021.108684

Aziz, M. A., Adnan, M., Khan, A. H., Rehman, A. U., Jan, R., & Khan, J. (2016). Ethno-medicinal survey of important plants practiced by indigenous community at Ladha subdivision, South Waziristan agency, Pakistan. *Journal of Ethnobiology and Ethnomedicine*, *12*(1), 53. https://doi.org/10.1186/s13002-016-0126-7

Balde, M. D., Soumah, A. M., Diallo, A., Sall, A. O., Mochache, V., Ahmed, W., Toure, A. O., Diallo, R., Camara, S., O'Neill, S., & Pallitto, C. C. (2022). Involving the health sector in the prevention and care of female genital mutilation: results from formative research in Guinea. *Reproductive Health*, *19*(1), 156. https://doi.org/10.1186/s12978-022-01428-4

Balogh, R., Whitelaw, S., & Thompson, J. (2008). Rapid Needs Appraisal in the modern NHS: potential and dilemmas. *Critical Public Health*, *18*(2), 233–244. https://doi.org/10.1080/09581590701377010

Barden-O'Fallon, J., Barry, M. A., Brodish, P., & Hazerjian, J. (2015). Rapid Assessment of Ebola-Related Implications for Reproductive, Maternal, Newborn and Child Health Service Delivery and Utilization in Guinea. *PLoS Currents*, *7*. https://doi.org/10.1371/currents.outbreaks.0b0ba06009dd091bc39ddb3c6d7b0826

Barreras, J. L., Linnemayr, S. L., & MacCarthy, S. (2019). 'We have a stronger survival mode': exploring knowledge gaps and culturally sensitive messaging of PrEP among Latino men who have sex with men and Latina transgender women in Los Angeles, CA. *AIDS Care*, *31*(10), 1221–1227. https://doi.org/10.1080/09540121.2019.1601669

Bayleyegn, T., Wolkin, A., Oberst, K., Young, S., Sanchez, C., Phelps, A., Schulte, J., Rubin, C., & Batts, D. (2006). Rapid assessment of the needs and health status in Santa Rosa and Escambia counties, Florida, after Hurricane Ivan, September 2004. *Disaster Management & Response : DMR : An Official Publication of the Emergency Nurses Association*, *4*(1), 12–18. https://doi.org/10.1016/j.dmr.2005.10.001

Beckerleg, S., Telfer, M., & Sadiq, A. (2006). A rapid assessment of heroin use in Mombasa, Kenya. *Substance Use & Misuse*, *41*(6–7), 1029–1044. https://doi.org/10.1080/10826080600667193

Belackova, V., Janikova, B., Vacek, J., Fidesova, H., & Miovsky, M. (2017). 'It can't happen to me': Alcohol drinkers on the 2012 outbreak of methanol poisonings and the subsequent prohibition in the Czech Republic. *Nordisk Alkohol- & Narkotikatidskrift : NAT*, *34*(5), 385–399. https://doi.org/10.1177/1455072517733597

Belford, M., Robertson, T., & Jepson, R. (2017). Using evaluability assessment to assess local community development health programmes: a Scottish case-study. *BMC Medical Research Methodology*, *17*(1), 70. https://doi.org/10.1186/s12874-017-0334-4

Betancourt, T. S., Speelman, L., Onyango, G., & Bolton, P. (2009). A qualitative study of mental health problems among children displaced by war in northern Uganda. *Transcultural Psychiatry*, *46*(2), 238–256. https://doi.org/10.1177/1363461509105815

Bjørkhaug, I., & Hatløy, A. (2009). Utilization of respondent-driven sampling among a population of child workers in the diamond-mining sector of Sierra Leone. *Global Public Health*, *4*(1), 96–109. https://doi.org/10.1080/17441690701464207

Brittain, A. W., August, E. M., Romero, L., Sheahan, M., Krashin, J., Ntansah, C., Honein, M. A., Jamieson, D. J., Ellis, E. M., Davis, M. S., & Lathrop, E. (2019). Community Perspectives on Contraception in the Context of the Zika Virus in the U.S. Virgin Islands: Implications for Communication and Messaging. *Women's Health Issues : Official Publication of the Jacobs Institute of Women's Health*, *29*(3), 245–251. https://doi.org/10.1016/j.whi.2019.01.007

Brown, D. R., Hernández, A., Saint-Jean, G., Evans, S., Tafari, I., Brewster, L. G., Celestin, M. J., Gómez-Estefan, C., Regalado, F., Akal, S., Nierenberg, B., Kauschinger, E. D., Schwartz, R., & Page, J. B. (2008). A participatory action research pilot study of urban health disparities using rapid assessment response and evaluation. *American Journal of Public Health*, *98*(1), 28–38. https://doi.org/10.2105/AJPH.2006.091363

Burgess-Allen, J., & Owen-Smith, V. (2010). Using mind mapping techniques for rapid qualitative data analysis in public participation processes. *Health Expectations : An International Journal of Public Participation in Health Care and Health Policy*, *13*(4), 406–415. https://doi.org/10.1111/j.1369-7625.2010.00594.x

Burks, D. J., Robbins, R., & Durtschi, J. P. (2011). American Indian gay, bisexual and two-spirit men: a rapid assessment of HIV/AIDS risk factors, barriers to prevention and culturally-sensitive intervention. *Culture, Health & Sexuality*, *13*(3), 283–298. https://doi.org/10.1080/13691058.2010.525666

Butler, J. R. A., Davila, F., Alders, R., Bourke, R. M., Crimp, S., McCarthy, J., McWilliam, A., Palo, A. S. M., Robins, L., Webb, M. J., van Wensveen, M., Sanderson, T., & Walker, D. (2021). A rapid assessment framework for food system shocks: Lessons learned from COVID-19 in the Indo-Pacific region. *Environmental Science & Policy*, *117*, 34–45. https://doi.org/10.1016/j.envsci.2020.12.011

Carter-Edwards, L., Grewe, M. E., Fair, A. M., Jenkins, C., Ray, N. J., Bilheimer, A., Dave, G., Nunez-Smith, M., Richmond, A., & Wilkins, C. H. (2021). Recognizing Cross-Institutional Fiscal and Administrative Barriers and Facilitators to Conducting Community-Engaged Clinical and Translational Research. *Academic Medicine : Journal of the Association of American Medical Colleges*, *96*(4), 558–567. https://doi.org/10.1097/ACM.0000000000003893

Chilanga, E., Dzimbiri, M., Mwanjawala, P., Keller, A., & Mbeya, R. A. (2022). Religion, politics and COVID-19 risk perception among urban residents in Malawi. *BMC Public Health*, *22*(1), 1430. https://doi.org/10.1186/s12889-022-13858-7

Chin, N. P., Goepp, J. G., Malia, T., & Poordabbagh, A. (2004). Planning Emergency Medical Services for Children in Bolivia. *Pediatric Emergency Care*, *20*(9), 593–598. https://doi.org/10.1097/01.pec.0000139737.08474.b0

Chopra, M., Doherty, T., Mehatru, S., & Tomlinson, M. (2009). Rapid assessment of infant feeding support to HIV-positive women accessing prevention of mother-to-child transmission services in Kenya, Malawi and Zambia. *Public Health Nutrition*, *12*(12), 2323–2328. https://doi.org/10.1017/S1368980009005606

Chopra, M., & Rollins, N. (2008). Infant feeding in the time of HIV: rapid assessment of infant feeding policy and programmes in four African countries scaling up prevention of mother to child transmission programmes. *Archives of Disease in Childhood*, *93*(4), 288–291. https://doi.org/10.1136/adc.2006.096321

Chung, A. D., Kwan, B. Y. M., Wagner, N., Braund, H., Hanmore, T., Hall, A. K., McEwan, L., Dalgarno, N., & Dagnone, J. D. (2022). An adaptation-focused evaluation of Canada's first competency-based medical education implementation in radiology. *European Journal of Radiology*, *147*, 110109. https://doi.org/10.1016/j.ejrad.2021.110109

Chung, J. W. Y., Wong, T. K. S., Chang, K. K. P., Chow, C. B., Chung, B. P. M., Chung, G., Ho, S., Ho, J. S. C., Lai, C. K. Y., Lai, A., Lam, V. S. F., Lau, J., Liu, J., Mok, E., & Wong, D. (2004). Rapid assessment of a helpdesk service supporting severe acute respiratory syndrome patients and their relatives. *Journal of Clinical Nursing*, *13*(6), 748–755. https://doi.org/10.1111/j.1365-2702.2004.00954.x

Cohn, W. F., Canan, C. E., Knight, S., Waldman, A. L., Dillingham, R., Ingersoll, K., Schexnayder, J., & Flickinger, T. E. (2021). An Implementation Strategy to Expand Mobile Health Use in HIV Care Settings: Rapid Evaluation Study Using the Consolidated Framework for Implementation Research. *JMIR MHealth and UHealth*, *9*(4), e19163. https://doi.org/10.2196/19163

Corcorran, M. A., Austin, E. J., Behrends, C. N., Briggs, E. S., Frost, M. C., Juarez, A. M., Frank, N. D., Healy, E., Prohaska, S. M., LaKosky, P. A., Kapadia, S. N., Perlman, D. C., Schackman, B. R., Des Jarlais, D. C., Williams, E. C., & Glick, S. N. (2023). Syringe Service Program Perspectives on Barriers, Readiness, and Programmatic Needs to Support Rollout of the COVID-19 Vaccine. *Journal of Addiction Medicine*, *17*(1), e36–e41. https://doi.org/10.1097/ADM.0000000000001036

Croucher, R., & Sohanpal, R. (2006). Improving access to dental care in East London's ethnic minority groups: community based, qualitative study. *Community Dental Health*, *23*(2), 95–100.

Dainty, K. N., & Kiran, T. (2020). 'Spending the day with your Family Health Team': rapid ethnography of a patient-centred quality improvement event. *BJGP Open*, *4*(1). https://doi.org/10.3399/bjgpopen20X101002

Dasgupta, R., Chaturvedi, S., Adhish, S. V., Ganguly, K. K., Rai, S., Sushant, L., & Arora, N. K. (2008). Social determinants and polio 'endgame': a qualitative study in high risk districts of India. *Indian Pediatrics*, *45*(5), 357–365.

Delobelle, P. A., Abbas, M., Datay, I., De Sa, A., Levitt, N., Schouw, D., & Reid, S. (2022). Non-communicable disease care and management in two sites of the Cape Town Metro during the first wave of COVID-19: A rapid appraisal. *African Journal of Primary Health Care & Family Medicine*, *14*(1), e1–e7. https://doi.org/10.4102/phcfm.v14i1.3215

Desmond, N., Allen, C. F., Clift, S., Justine, B., Mzugu, J., Plummer, M. L., Watson-Jones, D., & Ross, D. A. (2005). A typology of groups at risk of HIV/STI in a gold mining town in north-western Tanzania. *Social Science & Medicine*, *60*(8), 1739–1749. https://doi.org/10.1016/j.socscimed.2004.08.027

Doherty, T., Besada, D., Goga, A., Daviaud, E., Rohde, S., & Raphaely, N. (2017). 'If donors woke up tomorrow and said we can't fund you, what would we do?' A health system dynamics analysis of implementation of PMTCT option B+ in Uganda. *Globalization and Health*, *13*(1), 51. https://doi.org/10.1186/s12992-017-0272-2

Dos Santos, M. M., Trautmann, F., & Kools, J.-P. (2011). Rapid assessment response (RAR) study: drug use and health risk - Pretoria, South Africa. *Harm Reduction Journal*, *8*, 14. https://doi.org/10.1186/1477-7517-8-14

Douglass, K., Narayan, L., Allen, R., Pandya, J., & Talib, Z. (2021). Language diversity and challenges to communication in Indian emergency departments. *International Journal of Emergency Medicine*, *14*(1), 57. https://doi.org/10.1186/s12245-021-00380-7

Dozier, A. M., Block, R., Levy, D., Dye, T. D., & Pearson, T. A. (2008). Cardiovascular Health in the Developing World: Community Perceptions from Carriacou, Grenada. *CVD Prevention and Control*, *3*(3), 123–131. https://doi.org/10.1016/j.cvdpc.2008.05.001

Dozier, A. M., Ossip, D. J., Diaz, S., Sierra-Torres, E., Quiñones de Monegro, Z., Armstrong, L., Chin, N. P., & McIntosh, S. (2009). Health care workers in the Dominican Republic: self-perceived role in smoking cessation. *Evaluation & the Health Professions*, *32*(2), 144–164. https://doi.org/10.1177/0163278709333152

Dwyer, R., Power, R., Denham, G., & Dietze, P. (2014). Public injecting and public amenity in an inner-city suburb of Melbourne, Australia. *Journal of Substance Use*, 162–169. https://doi.org/10.3109/14659891.2014.987834

Ezard, N., Oppenheimer, E., Burton, A., Schilperoord, M., Macdonald, D., Adelekan, M., Sakarati, A., & van Ommeren, M. (2011). Six rapid assessments of alcohol and other substance use in populations displaced by conflict. *Conflict and Health*, *5*(1), 1. https://doi.org/10.1186/1752-1505-5-1

Fadholah, A., Istikomah, S. A., Islamanda, C. S., & Jannah, E. R. M. (2021). Identification of herbal products used by families in the campus of Darussalam Gontor University. *Pharmacy Education*, *21*(2), 31–35. https://doi.org/10.46542/pe.2021.212.3135

Forber-Pratt, A. J., Burdick, C. E., & Narasimham, G. (2022). Perspectives about COVID-19 vaccination among the paralysis community in the United States. *Rehabilitation Psychology*, *67*(1), 9–19. https://doi.org/10.1037/rep0000426

Foreman-Mackey, A., Bayoumi, A. M., Miskovic, M., Kolla, G., & Strike, C. (2019). 'It's our safe sanctuary': Experiences of using an unsanctioned overdose prevention site in Toronto, Ontario. *The International Journal on Drug Policy*, *73*, 135–140. https://doi.org/10.1016/j.drugpo.2019.09.019

Gaiha, S. M., Gulfam, F. R., Siddiqui, I., Kishore, R., & Krishnan, S. (2021). Pilot Community Mental Health Awareness Campaign Improves Service Coverage in India. *Community Mental Health Journal*, *57*(5), 814–827. https://doi.org/10.1007/s10597-020-00714-4

Gao, J., Zheng, P., Jia, Y., Chen, H., Mao, Y., Chen, S., Wang, Y., Fu, H., & Dai, J. (2020). Mental health problems and social media exposure during COVID-19 outbreak. *PLOS ONE*, *15*(4), e0231924. https://doi.org/10.1371/journal.pone.0231924

Gawaya, M., Terrill, D., & Williams, E. (2022). Using rapid evaluation methods to assess service delivery changes: Lessons learned for evaluation practice during the COVID-19 pandemic. *Evaluation Journal of Australasia*, *22*(1), 30–48. https://doi.org/10.1177/1035719X211057630

Goepp, J., Chin, N. P., Malia, T., & Poordabbagh, A. (2004). Planning Emergency Medical Services for Children in Bolivia. *Pediatric Emergency Care*, *20*(10), 664–670. https://doi.org/10.1097/01.pec.0000142950.58265.15

Grant, L., Brown, J., Leng, M., Bettega, N., & Murray, S. A. (2011). Palliative care making a difference in rural Uganda, Kenya and Malawi: three rapid evaluation field studies. *BMC Palliative Care*, *10*, 8. https://doi.org/10.1186/1472-684X-10-8

Green, T. C., Bowman, S. E., Ray, M., Zaller, N., Heimer, R., & Case, P. (2013). Collaboration or coercion? Partnering to divert prescription opioid medications. *Journal of Urban Health : Bulletin of the New York Academy of Medicine*, *90*(4), 758–767. https://doi.org/10.1007/s11524-012-9784-5

Guise, A., Witzel, T. C., Mandal, S., Sabin, C., Rhodes, T., Nardone, A., & Harris, M. (2018). A qualitative assessment of the acceptability of hepatitis C remote self-testing and self-sampling amongst people who use drugs in London, UK. *BMC Infectious Diseases*, *18*(1), 281. https://doi.org/10.1186/s12879-018-3185-7

Hammond, N., Steels, S., & King, G. (2022). Contraceptive and pregnancy concerns in the UK during the first COVID-19 lockdown: A rapid study. *Sexual & Reproductive Healthcare : Official Journal of the Swedish Association of Midwives*, *33*, 100754. https://doi.org/10.1016/j.srhc.2022.100754

Hanvoravongchai, P., Adisasmito, W., Chau, P. N., Conseil, A., de Sa, J., Krumkamp, R., Mounier-Jack, S., Phommasack, B., Putthasri, W., Shih, C.-S., Touch, S., Coker, R., & AsiaFluCap Project. (2010). Pandemic influenza preparedness and health systems challenges in Asia: results from rapid analyses in 6 Asian countries. *BMC Public Health*, *10*, 322. https://doi.org/10.1186/1471-2458-10-322

Harapan, H., Alleta, A., Anwar, S., Setiawan, A. M., Maulana, R., Wahyuniati, N., Ramadana, M. R., Ikram, I., Haryanto, S., Jamil, K. F., Kuch, U., & Rodríguez-Morales, A. J. (2018). Attitudes towards Zika virus infection among medical doctors in Aceh province, Indonesia. *Journal of Infection and Public Health*, *11*(1), 99–104. https://doi.org/10.1016/j.jiph.2017.06.013

Hawkins, K., Price, N., & Mussa, F. (2009). Milking the cow: young women's construction of identity and risk in age-disparate transactional sexual relationships in Maputo, Mozambique. *Global Public Health*, *4*(2), 169–182. https://doi.org/10.1080/17441690701589813

Hipgrave, D. B., Anderson, I., & Sato, M. (2019). A rapid assessment of the political economy of health at district level, with a focus on maternal, newborn and child health, in Bangladesh, Indonesia, Nepal and the Philippines. *Health Policy and Planning*, *34*(10), 762–772. https://doi.org/10.1093/heapol/czz082

Holdsworth, L. M., Safaeinili, N., Winget, M., Lorenz, K. A., Lough, M., Asch, S., & Malcolm, E. (2020). Adapting rapid assessment procedures for implementation research using a team-based approach to analysis: a case example of patient quality and safety interventions in the ICU. *Implementation Science*, *15*(1), 12. https://doi.org/10.1186/s13012-020-0972-5

Hopkinson, B., Balabanova, D., McKee, M., & Kutzin, J. (2004). The human perspective on health care reform: coping with diabetes in Kyrgyzstan. *The International Journal of Health Planning and Management*, *19*(1), 43–61. https://doi.org/10.1002/hpm.745

Hossain, S. M. M., & Kolsteren, P. (2003). The 1998 Flood in Bangladesh: Is Different Targeting Needed During Emergencies and Recovery to Tackle Malnutrition? *Disasters*, *27*(2), 172–184. https://doi.org/10.1111/1467-7717.00227

Jack, E. (2020). Service user involvement in an undergraduate nursing programme. *The Journal of Mental Health Training, Education and Practice*, *15*(3), 125–140. https://doi.org/10.1108/JMHTEP-12-2018-0073

Joarder, T., Cooper, A., & Zaman, S. (2014). Meaning of death: an exploration of perception of elderly in a Bangladeshi village. *Journal of Cross-Cultural Gerontology*, *29*(3), 299–314. https://doi.org/10.1007/s10823-014-9237-6

Jumbe, S., Milner, A., Clinch, M., Kennedy, J., Pinder, R. J., Sharpe, C. A., & Fenton, K. (2021). A qualitative evaluation of Southwark Council's public health response to mitigating the mental health impact of the 2017 London bridge and borough market terror attack. *BMC Public Health*, *21*(1), 1427. https://doi.org/10.1186/s12889-021-11447-8

Kahle, E. M., Barash, E. A., Page, L. C., Lansky, A., Jafa, K., Sullivan, P. S., & Buskin, S. E. (2009). Evaluation of the impact of news coverage of an HIV multiclass drug-resistant cluster in Seattle, Washington. *American Journal of Public Health*, *99 Suppl 1*(Suppl 1), S131-6. https://doi.org/10.2105/AJPH.2007.126656

Kalil, F. S., Bedaso, M. H., Abdulle, M. S., & Mohammed, N. U. (2021). Evaluation of Measles Surveillance Systems in Ginnir District, Bale Zone, Southeast Ethiopia: A Concurrent Embedded Mixed Quantitative/Qualitative Study. *Risk Management and Healthcare Policy*, *14*, 997–1008. https://doi.org/10.2147/RMHP.S295889

Kamineni, V. V., Turk, T., Wilson, N., Satyanarayana, S., & Chauhan, L. S. (2011). A rapid assessment and response approach to review and enhance advocacy, communication and social mobilisation for tuberculosis control in Odisha state, India. *BMC Public Health*, *11*, 463. https://doi.org/10.1186/1471-2458-11-463

Kiawi, E., McLellan-Lemal, E., Mosoko, J., Chillag, K., & Raghunathan, P. L. (2012). ‘Research participants want to feel they are better off than they were before research was introduced to them’: engaging cameroonian rural plantation populations in HIV research. *BMC International Health and Human Rights*, *12*, 8. https://doi.org/10.1186/1472-698X-12-8

Kibe, L. W., Habluetzel, A., Gachigi, J. K., Kamau, A. W., & Mbogo, C. M. (2019). Exploring communities’ and health workers’ perceptions of indicators and drivers of malaria decline in Malindi, Kenya. *MalariaWorld Journal*, *8*, 21.

Kodish, S., Aburto, N., Dibari, F., Brieger, W., Agostinho, S. P., & Gittelsohn, J. (2015). Informing a Behavior Change Communication Strategy: Formative Research Findings From the Scaling Up Nutrition Movement in Mozambique. *Food and Nutrition Bulletin*, *36*(3), 354–370. https://doi.org/10.1177/0379572115598447

Kodish, S., Aburto, N., Hambayi, M. N., Kennedy, C., & Gittelsohn, J. (2015). Identifying the Sociocultural Barriers and Facilitating Factors to Nutrition-related Behavior Change: Formative Research for a Stunting Prevention Program in Ntchisi, Malawi. *Food and Nutrition Bulletin*, *36*(2), 138–153. https://doi.org/10.1177/0379572115586784

Kraaij-Dirkzwager, M., van der Ree, J., & Lebret, E. (2017). Rapid Assessment of Stakeholder Concerns about Public Health. An Introduction to a Fast and Inexpensive Approach Applied on Health Concerns about Intensive Animal Production Systems. *International Journal of Environmental Research and Public Health*, *14*(12). https://doi.org/10.3390/ijerph14121534

Kumar, M., Macharia, P., Nyongesa, V., Kathono, J., Yator, O., Mwaniga, S., McKay, M., Huang, K. Y., Shidhaye, R., Njuguna, S., & Saxena, S. (2022). Human-centered design exploration with Kenyan health workers on proposed digital mental health screening and intervention training development: Thematic analysis of user preferences and needs. *DIGITAL HEALTH*, *8*, 205520762210900. https://doi.org/10.1177/20552076221090035

Kumar, R., Shaikh, B. T., Ahmed, J., Khan, Z., Mursalin, S., Memon, M. I., & Zareen, S. (2013). The human resource information system: a rapid appraisal of Pakistan’s capacity to employ the tool. *BMC Medical Informatics and Decision Making*, *13*, 104. https://doi.org/10.1186/1472-6947-13-104

Kyamwanga, I. T., Turyakira, E., Kilbourne-Brook, M., & Coffey, P. S. (2014). Potential for revitalisation of the diaphragm for family planning in Uganda: a rapid assessment of the feasibility of introducing the SILCS Diaphragm. *African Journal of Reproductive Health*, *18*(2), 77–86.

Laisser, R., Danna, V. A., Bonet, M., Oladapo, O. T., & Lavender, T. (2021). An exploration of midwives' views of the latest World Health Organization labour care guide. *African Journal of Midwifery and Women's Health*, *15*(4), 1–11. https://doi.org/10.12968/ajmw.2020.0043

Leonard, A. L., Verster, A., & Coetzee, M. (2014). Developing family-friendly signage in a South African paediatric healthcare setting. *Curationis*, *37*(2), 1–7. https://doi.org/10.4102/curationis.v37i2.1250

Lim, S. C., Kataria, I., Ngongo, C., Usek, V. S., Kudtarkar, S. R., Chandran, A., & Mustapha, F. I. (2022). Exploring the impact of COVID-19 movement control orders on eating habits and physical activity in low-resource urban settings in Malaysia. *Global Health Promotion*, *29*(4), 17579759221091196. https://doi.org/10.1177/17579759221091197

Liu, Y., Dao, Z., Yang, C., Liu, Y., & Long, C. (2009). Medicinal plants used by Tibetans in Shangri-la, Yunnan, China. *Journal of Ethnobiology and Ethnomedicine*, *5*, 15. https://doi.org/10.1186/1746-4269-5-15

Livorsi, D., Knobloch, M. J., Blue, L. A., Swafford, K., Maze, L., Riggins, K., Hayward, T., & Safdar, N. (2016). A rapid assessment of barriers and facilitators to safety culture in an intensive care unit. *International Nursing Review*, *63*(3), 372–376. https://doi.org/10.1111/inr.12254

Logez, S., Soyolgerel, G., Fields, R., Luby, S., & Hutin, Y. (2004). Rapid assessment of injection practices in Mongolia. *American Journal of Infection Control*, *32*(1), 31–37. https://doi.org/10.1016/j.ajic.2003.06.006

Loko, L. E. Y., Medegan Fagla, S., Orobiyi, A., Glinma, B., Toffa, J., Koukoui, O., Djogbenou, L., & Gbaguidi, F. (2019). Traditional knowledge of invertebrates used for medicine and magical–religious purposes by traditional healers and indigenous populations in the Plateau Department, Republic of Benin. *Journal of Ethnobiology and Ethnomedicine*, *15*(1), 66. https://doi.org/10.1186/s13002-019-0344-x

Luz, T. C. B., Tavares, N. U. L., de Castro, A. K. S., Marques, I. C., Dos Santos, E. M., & Cota, B. B. (2022). MedMinas project: design and use of mixed methods in the evaluation of pharmaceutical services in primary health care in Minas Gerais, Brazil. *BMC Medical Research Methodology*, *22*(1), 80. https://doi.org/10.1186/s12874-022-01568-y

Lynch, K. A., Omisore, A. D., Famurewa, O., Olasehinde, O., Odujoko, O., Vera, J., Kingham, T. P., Alatise, O. I., Egberongbe, A. A., Morris, E. A., Atkinson, T. M., & Sutton, E. J. (2021). Designing Participatory Needs Assessments to Support Global Health Interventions in Time-Limited Settings: A Case Study From Nigeria. *International Journal of Qualitative Methods*, *20*. https://doi.org/10.1177/16094069211002421

Maalim, A. D. (2006). Participatory rural appraisal techniques in disenfranchised communities: a Kenyan case study. *International Nursing Review*, *53*(3), 178–188. https://doi.org/10.1111/j.1466-7657.2006.00489.x

Mahmood, A., Mahmood, A., Malik, R. N., & Shinwari, Z. K. (2013). Indigenous knowledge of medicinal plants from Gujranwala district, Pakistan. *Journal of Ethnopharmacology*, *148*(2), 714–723. https://doi.org/10.1016/j.jep.2013.05.035

Mahmood, A., Rashid, S., & Malik, R. N. (2013). Determination of toxic heavy metals in indigenous medicinal plants used in Rawalpindi and Islamabad cities, Pakistan. *Journal of Ethnopharmacology*, *148*(1), 158–164. https://doi.org/10.1016/j.jep.2013.04.005

Maroyi, A. (2011). An ethnobotanical survey of medicinal plants used by the people in Nhema communal area, Zimbabwe. *Journal of Ethnopharmacology*, *136*(2), 347–354. https://doi.org/10.1016/j.jep.2011.05.003

Miller, S., Cordero, M., Coleman, A. L., Figueroa, J., Brito-Anderson, S., Dabagh, R., Calderon, V., Cáceres, F., Fernandez, A. J., & Nunez, M. (2003). Quality of care in institutionalized deliveries: the paradox of the Dominican Republic. *International Journal of Gynecology & Obstetrics*, *82*(1), 89–103. https://doi.org/10.1016/S0020-7292(03)00148-6

Mital, S., Miles, G., McLellan-Lemal, E., Muthui, M., & Needle, R. (2016). Heroin shortage in Coastal Kenya: A rapid assessment and qualitative analysis of heroin users' experiences. *The International Journal on Drug Policy*, *30*, 91–98. https://doi.org/10.1016/j.drugpo.2015.08.010

Mitchinson, L., Dowrick, A., Buck, C., Hoernke, K., Martin, S., Vanderslott, S., Robinson, H., Rankl, F., Manby, L., Lewis-Jackson, S., & Vindrola-Padros, C. (2021). Missing the human connection: A rapid appraisal of healthcare workers' perceptions and experiences of providing palliative care during the COVID-19 pandemic. *Palliative Medicine*, *35*(5), 852–861. https://doi.org/10.1177/02692163211004228

Moloney, K., Scheuer, H., Engstrom, A., Schreiber, M., Whiteside, L., Nehra, D., Walen, M. Lou, Rivara, F., & Zatzick, D. (2020). Experiences and Insights from the Early US COVID-19 Epicenter: A Rapid Assessment Procedure Informed Clinical Ethnography Case Series. *Psychiatry*, *83*(2), 115–127. https://doi.org/10.1080/00332747.2020.1750214

Moodie, S. M., Tsui, E. K., & Silbergeld, E. K. (2010). Community- and family-level factors influence care-giver choice to screen blood lead levels of children in a mining community. *Environmental Research*, *110*(5), 484–496. https://doi.org/10.1016/j.envres.2010.03.012

Morin, S. F., Morfit, S., Maiorana, A., Aramrattana, A., Goicochea, P., Mutsambi, J. M., Robbins, J. L., & Richards, T. A. (2008). Building community partnerships: case studies of Community Advisory Boards at research sites in Peru, Zimbabwe, and Thailand. *Clinical Trials (London, England)*, *5*(2), 147–156. https://doi.org/10.1177/1740774508090211

Morojele, N. K., Kachieng'a, M. A., Mokoko, E., Nkoko, M. A., Parry, C. D. H., Nkowane, A. M., Moshia, K. M., & Saxena, S. (2006). Alcohol use and sexual behaviour among risky drinkers and bar and shebeen patrons in Gauteng province, South Africa. *Social Science & Medicine (1982)*, *62*(1), 217–227. https://doi.org/10.1016/j.socscimed.2005.05.031

Mueller, J. G., Assanou, I. H. B., Dan Guimbo, I., & Almedom, A. M. (2010). Evaluating rapid participatory rural appraisal as an assessment of ethnoecological knowledge and local biodiversity patterns. *Conservation Biology : The Journal of the Society for Conservation Biology*, *24*(1), 140–150. https://doi.org/10.1111/j.1523-1739.2009.01392.x

Mullane, S. L., Connolly, D., & Buman, M. P. (2019). The Perceived Value of Reducing Sedentary Behavior in the Truck Driving Population. *Frontiers in Public Health*, *7*, 214. https://doi.org/10.3389/fpubh.2019.00214

Murphy, A., Chikovani, I., Uchaneishvili, M., Makhashvili, N., & Roberts, B. (2018). Barriers to mental health care utilization among internally displaced persons in the republic of Georgia: a rapid appraisal study. *BMC Health Services Research*, *18*(1), 306. https://doi.org/10.1186/s12913-018-3113-y

Myers, J. J., Maiorana, A., Chapman, K., Lall, R., Kassie, N., & Persaud, N. (2011). How the Illicit Drug Economy Contributes to HIV Risk in St Vincent and the Grenadines. *Journal of the International Association of Physicians in AIDS Care (Chicago, Ill. : 2002)*, *10*(6), 396–406. https://doi.org/10.1177/1545109711418508

Naylor, G., Burke, L. A., & Holman, J. A. (2020). Covid-19 Lockdown Affects Hearing Disability and Handicap in Diverse Ways: A Rapid Online Survey Study. *Ear and Hearing*, *41*(6), 1442–1449. https://doi.org/10.1097/AUD.0000000000000948

Neal, P., Knowles, A., & DuMond, S. (2018). Fostering Community Engagement and Acquiring Understanding of Health Needs through a Participatory Rural Appraisal in Haiti. *Progress in*

*Community Health Partnerships : Research, Education, and Action*, *12*(4), 389–394. https://doi.org/10.1353/cpr.2018.0064
Needle, R., Kroeger, K., Belani, H., Achrekar, A., Parry, C. D., & Dewing, S. (2008). Sex, drugs, and HIV: rapid assessment of HIV risk behaviors among street-based drug using sex workers in Durban, South Africa. *Social Science & Medicine (1982)*, *67*(9), 1447–1455. https://doi.org/10.1016/j.socscimed.2008.06.031
Negandhi, H., Tiwari, R., Sharma, A., Nair, R., Zodpey, S., Reddy Allam, R., & Oruganti, G. (2017). Rapid assessment of facilitators and barriers related to the acceptance, challenges and community perception of daily regimen for treating tuberculosis in India. *Global Health Action*, *10*(1), 1290315. https://doi.org/10.1080/16549716.2017.1290315
Nemser, B., Aung, K., Mushamba, M., Chirwa, S., Sera, D., Chikhwaza, O., & Kachale, F. (2018). Data-informed decision-making for life-saving commodities investments in Malawi: A qualitative case study. *Malawi Medical Journal : The Journal of Medical Association of Malawi*, *30*(2), 111–119. https://doi.org/10.4314/mmj.v30i2.11
Nsibande, D., Loveday, M., Daniels, K., Sanders, D., Doherty, T., & Zembe, W. (2018). Approaches and strategies used in the training and supervision of Health Extension Workers (HEWs) delivering integrated community case management (iCCM) of childhood illness in Ethiopia: a qualitative rapid appraisal. *African Health Sciences*, *18*(1), 188–197. https://doi.org/10.4314/ahs.v18i1.24
Okuthe, O. S., McLeod, A., Otte, J. M., & Buyu, G. E. (2003). Use of rapid rural appraisal and cross-sectional studies in the assessment of constraints in smallholder cattle production systems in the western Kenya highlands. *The Onderstepoort Journal of Veterinary Research*, *70*(3), 237–242.
Oloukoi, G., Bob, U., & Jaggernath, J. (2014). Perception and trends of associated health risks with seasonal climate variation in Oke-Ogun region, Nigeria. *Health & Place*, *25*, 47–55. https://doi.org/10.1016/j.healthplace.2013.09.009
O'Meara, L., Turner, C., Coitinho, D. C., & Oenema, S. (2022). Consumer experiences of food environments during the Covid-19 pandemic: Global insights from a rapid online survey of individuals from 119 countries. *Global Food Security*, *32*, 100594. https://doi.org/10.1016/j.gfs.2021.100594
Palinkas, L. A., Engstrom, A., Whiteside, L., Moloney, K., & Zatzick, D. (2022). A Rapid Ethnographic Assessment of the Impact of the COVID-19 Pandemic on Mental Health Services Delivery in an Acute Care Medical Emergency Department and Trauma Center. *Administration and Policy in Mental Health*, *49*(2), 157–167. https://doi.org/10.1007/s10488-021-01154-2
Palinkas, L. A., Prussing, E., Reznik, V. M., & Landsverk, J. A. (2004). The San Diego East County school shootings: a qualitative study of community-level post-traumatic stress. *Prehospital and Disaster Medicine*, *19*(1), 113–121. https://doi.org/10.1017/s1049023x00001564
Palinkas, L. A., Springgate, B. F., Sugarman, O. K., Hancock, J., Wennerstrom, A., Haywood, C., Meyers, D., Johnson, A., Polk, M., Pesson, C. L., Seay, J. E., Stallard, C. N., & Wells, K. B. (2021). A Rapid Assessment of Disaster Preparedness Needs and Resources during the COVID-19 Pandemic. *International Journal of Environmental Research and Public Health*, *18*(2). https://doi.org/10.3390/ijerph18020425
Parry, C., Petersen, P., Dewing, S., Carney, T., Needle, R., Kroeger, K., & Treger, L. (2008). Rapid assessment of drug-related HIV risk among men who have sex with men in three South African cities. *Drug and Alcohol Dependence*, *95*(1–2), 45–53. https://doi.org/10.1016/j.drugalcdep.2007.12.005

Peltzer, K., Phaswana-Mafuya, N., & Ladzani, R. (2010). Implementation of the national programme for prevention of mother-to-child transmission of HIV: a rapid assessment in Cacadu district, South Africa. *African Journal of AIDS Research : AJAR*, *9*(1), 95–106. https://doi.org/10.2989/16085906.2010.484594

Pepall, E., Earnest, J., & James, R. (2007). Understanding community perceptions of health and social needs in a rural Balinese village: results of a rapid participatory appraisal. *Health Promotion International*, *22*(1), 44–52. https://doi.org/10.1093/heapro/dal042

Polidoro, B. A., Dahlquist, R. M., Castillo, L. E., Morra, M. J., Somarriba, E., & Bosque-Pérez, N. A. (2008). Pesticide application practices, pest knowledge, and cost-benefits of plantain production in the Bribri-Cabécar Indigenous Territories, Costa Rica. *Environmental Research*, *108*(1), 98–106. https://doi.org/10.1016/j.envres.2008.04.003

Poteat, T., Diouf, D., Drame, F. M., Ndaw, M., Traore, C., Dhaliwal, M., Beyrer, C., & Baral, S. (2011). HIV risk among MSM in Senegal: a qualitative rapid assessment of the impact of enforcing laws that criminalize same sex practices. *PloS One*, *6*(12), e28760. https://doi.org/10.1371/journal.pone.0028760

Quach, T., Đoàn, L. N., Liou, J., & Ponce, N. A. (2021). A Rapid Assessment of the Impact of COVID-19 on Asian Americans: Cross-sectional Survey Study. *JMIR Public Health and Surveillance*, *7*(6), e23976. https://doi.org/10.2196/23976

Rahman, M. R., Faiz, M. A., Nu, M. Y., Hassan, M. R., Chakrabarty, A. K., Kabir, I., Islam, K., Jafarullah, A. K. M., Alakabawy, M., Khatami, A., & Rashid, H. (2020). A Rapid Assessment of Health Literacy and Health Status of Rohingya Refugees Living in Cox's Bazar, Bangladesh Following the August 2017 Exodus from Myanmar: A Cross-Sectional Study. *Tropical Medicine and Infectious Disease*, *5*(3). https://doi.org/10.3390/tropicalmed5030110

Rhodes, S. D., Yee, L. J., & Hergenrather, K. C. (2006). A community-based rapid assessment of HIV behavioural risk disparities within a large sample of gay men in southeastern USA: a comparison of African American, Latino and white men. *AIDS Care*, *18*(8), 1018–1024. https://doi.org/10.1080/09540120600568731

Rinehart, D. J., Stowell, M., Barrett, K., Langland, K., Thomas-Gale, T., Al-Tayyib, A., & O'Connell, R. (2023). Exploring Family Planning Perspectives Among Men Receiving Medications for Opioid Use Disorder: Implications for Service Development. *Journal of Addiction Medicine*, *17*(1), 21–27. https://doi.org/10.1097/ADM.0000000000001012

Robles, M. C., Corches, C. L., Bradford, M., Rice, T. S., Sukul, D., Springer, M. V, Bailey, S., Oliver, A., & Skolarus, L. E. (2021). Understanding and Informing Community Emergency Cardiovascular Disease Preparedness during the COVID-19 Pandemic: Stroke Ready. *Journal of Stroke and Cerebrovascular Diseases : The Official Journal of National Stroke Association*, *30*(2), 105479. https://doi.org/10.1016/j.jstrokecerebrovasdis.2020.105479

Romeu-Labayen, M., Tort-Nasarre, G., Alvarez, B., Subias-Miquel, M., Vázquez-Segura, E., Marre, D., & Galbany-Estragués, P. (2022). Spanish nurses' experiences with personal protective equipment and perceptions of risk of contagion from COVID-19: A qualitative rapid appraisal. *Journal of Clinical Nursing*, *31*(15–16), 2154–2166. https://doi.org/10.1111/jocn.16031

Sambo, E., Bettridge, J., Dessie, T., Amare, A., Habte, T., Wigley, P., & Christley, R. M. (2015). Participatory evaluation of chicken health and production constraints in Ethiopia. *Preventive Veterinary Medicine*, *118*(1), 117–127. https://doi.org/10.1016/j.prevetmed.2014.10.014

Seay, J. S., Carrasquillo, O., Campos, N. G., McCann, S., Amofah, A., Pierre, L., & Kobetz, E. (2015). Cancer Screening Utilization Among Immigrant Women in Miami, Florida. *Progress in*

*Community Health Partnerships : Research, Education, and Action*, *9 Suppl*, 11–20. https://doi.org/10.1353/cpr.2015.0029

Seidel, S., Muciimi, J., Chang, J., Gitari, S., Keiser, P., & Goodman, M. L. (2018). Community perceptions of home environments that lead children & youth to the street in semi-rural Kenya. *Child Abuse & Neglect*, *82*, 34–44. https://doi.org/10.1016/j.chiabu.2018.05.011

Shah, S., Rollins, N. C., Bland, R., & Child Health Group. (2005). Breastfeeding knowledge among health workers in rural South Africa. *Journal of Tropical Pediatrics*, *51*(1), 33–38. https://doi.org/10.1093/tropej/fmh071

Shamsuddin, M., Alam, M. M., Hossein, M. S., Goodger, W. J., Bari, F. Y., Ahmed, T. U., Hossain, M. M., & Khan, A. H. M. S. I. (2007). Participatory rural appraisal to identify needs and prospects of market-oriented dairy industries in Bangladesh. *Tropical Animal Health and Production*, *39*(8), 567–581. https://doi.org/10.1007/s11250-007-9062-9

Sharma, S. V., Haidar, A., Noyola, J., Tien, J., Rushing, M., Naylor, B. M., Chuang, R.-J., & Markham, C. (2020). Using a rapid assessment methodology to identify and address immediate needs among low-income households with children during COVID-19. *PLOS ONE*, *15*(10), e0240009. https://doi.org/10.1371/journal.pone.0240009

Shimkhada, R., Attai, D., Scheitler, A. J., Babey, S., Glenn, B., & Ponce, N. (2021). Using a Twitter Chat to Rapidly Identify Barriers and Policy Solutions for Metastatic Breast Cancer Care: Qualitative Study. *JMIR Public Health and Surveillance*, *7*(1), e23178. https://doi.org/10.2196/23178

Sithole, Z., Nyadzayo, T., Kanyowa, T., Mathieu, J., Kambarami, T., Nemaramba, M., Machaka, R., Bekele, H., Njovo, H., & Gasasira, A. (2021). Enhancing capacity of Zimbabwe's health system to respond to climate change induced drought: a rapid nutritional assessment. *The Pan African Medical Journal*, *40*, 113. https://doi.org/10.11604/pamj.2021.40.113.30545

Solomon, P. L., Tennille, J. A., Lipsitt, D., Plumb, E., Metzger, D., & Blank, M. B. (2007). Rapid assessment of existing HIV prevention programming in a community mental health center. *Journal of Prevention & Intervention in the Community*, *33*(1–2), 137–151. https://doi.org/10.1300/J005v33n01_11

Springgate, B. F., Allen, C., Jones, C., Lovera, S., Meyers, D., Campbell, L., Palinkas, L. A., & Wells, K. B. (2009). Rapid community participatory assessment of health care in post-storm New Orleans. *American Journal of Preventive Medicine*, *37*(6 Suppl 1), S237-43. https://doi.org/10.1016/j.amepre.2009.08.007

Stajduhar, K. I., Poffenroth, L., Wong, E., Archibald, C. P., Sutherland, D., & Rekart, M. (2004). Missed opportunities: injection drug use and HIV/AIDS in Victoria, Canada. *International Journal of Drug Policy*, *15*(3), 171–181. https://doi.org/10.1016/j.drugpo.2004.01.001

Sy, A., Marriott, J., Tannis, C., Demment, M., McIntosh, S., Hadley, J., Albert, P., Buenconsejo-Lum, L., & Dye, T. (2020). A Rapid Assessment Procedure to Develop A Non-Communicable Disease Prevention Pilot Health Communications Project Using E- and M-Health Communications in Pohnpei State, Federated States of Micronesia. *Hawai'i Journal of Health & Social Welfare*, *79*(6 Suppl 2), 58–63.

Theiss-Nyland, K., Ejersa, W., Karema, C., Koné, D., Koenker, H., Cyaka, Y., Lynch, M., Webster, J., & Lines, J. (2016). Operational challenges to continuous LLIN distribution: a qualitative rapid assessment in four countries. *Malaria Journal*, *15*, 131. https://doi.org/10.1186/s12936-016-1184-y

Tindana, P., Bull, S., Amenga-Etego, L., de Vries, J., Aborigo, R., Koram, K., Kwiatkowski, D., & Parker, M. (2012). Seeking consent to genetic and genomic research in a rural Ghanaian setting: a qualitative study of the MalariaGEN experience. *BMC Medical Ethics*, *13*, 15. https://doi.org/10.1186/1472-6939-13-15

Tort-Nasarre, G., Alvarez, B., Galbany-Estragués, P., Subías-Miquel, M., Vázquez-Segura, E., Marre, D., & Romeu-Labayen, M. (2021). Front-line nurses' responses to organisational changes during the COVID-19 in Spain: A qualitative rapid appraisal. *Journal of Nursing Management*, *29*(7), 1983–1991. https://doi.org/10.1111/jonm.13362

Tran, B. X., Hoang, C. L., Nguyen, N. T. T., Le, H. T., Pham, H. Q., Hoang, M. T., Nguyen, T. H., Latkin, C. A., Ho, C. S. H., & Ho, R. C. M. (2021). COVID-19 Preparedness and Response: Validation of a Rapid Assessment Tool to Evaluate Priorities of Health Workers at the Grassroots Level. *Frontiers in Public Health*, *9*, 562600. https://doi.org/10.3389/fpubh.2021.562600

Turk, T., Latu, N., Cocker-Palu, E., Liavaa, V., Vivili, P., Gloede, S., & Simons, A. (2013). Using rapid assessment and response to operationalise physical activity strategic health communication campaigns in Tonga. *Health Promotion Journal of Australia : Official Journal of Australian Association of Health Promotion Professionals*, *24*(1), 13–19. https://doi.org/10.1071/HE12903

Turk, T., Quang, N. D., Nga, T. T., Phuong, H., Tung, L. V. A., & Trang, V. H. (2017). A rapid assessment and response approach for socially marketed nutrition commodities in Viet Nam. *Asia Pacific Journal of Clinical Nutrition*, *26*(1), 182–189. https://doi.org/10.6133/apjcn.072016.12

Turkson, P. K. (2009). Client's satisfaction with delivery of animal health-care services in peri-urban Ghana. *Preventive Veterinary Medicine*, *90*(3–4), 153–159. https://doi.org/10.1016/j.prevetmed.2009.04.012

Utarini, A., Winkvist, A., & Ulfa, F. M. (2003). Rapid assessment procedures of malaria in low endemic countries: community perceptions in Jepara district, Indonesia. *Social Science & Medicine*, *56*(4), 701–712. https://doi.org/10.1016/S0277-9536(02)00066-7

van der Merwe, M., D'Ambruoso, L., Witter, S., Twine, R., Mabetha, D., Hove, J., Byass, P., Tollman, S., & Kahn, K. (2021). Collective reflections on the first cycle of a collaborative learning platform to strengthen rural primary healthcare in Mpumalanga, South Africa. *Health Research Policy and Systems*, *19*(1), 66. https://doi.org/10.1186/s12961-021-00716-y

Van Hout, M. C., & Bingham, T. (2013). Open drug scenes and drug-related public nuisance: a visual rapid assessment research study in Dublin, Ireland. *Journal of Ethnicity in Substance Abuse*, *12*(2), 154–178. https://doi.org/10.1080/15332640.2013.788917

van Kamp, I., van der Velden, P. G., Stellato, R. K., Roorda, J., van Loon, J., Kleber, R. J., Gersons, B. B. R., & Lebret, E. (2006). Physical and mental health shortly after a disaster: first results from the Enschede firework disaster study. *European Journal of Public Health*, *16*(3), 253–259. https://doi.org/10.1093/eurpub/cki188

Van Meer, R., Hohenadel, K., Fitzgerald-Husek, A., Warshawsky, B., Sider, D., Schwartz, B., & Nelder, M. P. (2017). Zika Virus in Ontario: Evaluating a Rapid Risk Assessment Tool for Emerging Infectious Disease Threats. *Health Security*, *15*(3), 230–237. https://doi.org/10.1089/hs.2016.0127

Vidal-Infer, A., Tomás-Dols, S., Aguilar-Moya, R., Samper-Gras, T., Zarza, M. J., & Aguilar-Serrano, J. (2009). Christmas work dinners. A pattern of recreational use of alcohol and other drugs? *Adicciones*, *21*(2), 133–142.

Vindrola-Padros, C., Chisnall, G., Cooper, S., Dowrick, A., Djellouli, N., Symmons, S. M., Martin, S., Singleton, G., Vanderslott, S., Vera, N., & Johnson, G. A. (2020). Carrying Out Rapid

Qualitative Research During a Pandemic: Emerging Lessons From COVID-19. *Qualitative Health Research*, *30*(14), 2192–2204. https://doi.org/10.1177/1049732320951526

von Thiele Schwarz, U., Andersson, K., & Loeb, C. (2021). Quick and dirty or rapid and informative? Exploring a participatory method to facilitate implementation research and organizational change. *Journal of Health Organization and Management*, *ahead-of-print*(ahead-of-print), 868–885. https://doi.org/10.1108/JHOM-12-2020-0503

Waiswa, P., Kemigisa, M., Kiguli, J., Naikoba, S., Pariyo, G. W., & Peterson, S. (2008). Acceptability of evidence-based neonatal care practices in rural Uganda - implications for programming. *BMC Pregnancy and Childbirth*, *8*, 21. https://doi.org/10.1186/1471-2393-8-21

Walton, H., Vindrola-Padros, C., Crellin, N. E., Sidhu, M. S., Herlitz, L., Litchfield, I., Ellins, J., Ng, P. L., Massou, E., Tomini, S. M., & Fulop, N. J. (2022). Patients' experiences of, and engagement with, remote home monitoring services for COVID-19 patients: A rapid mixed-methods study. *Health Expectations : An International Journal of Public Participation in Health Care and Health Policy*, *25*(5), 2386–2404. https://doi.org/10.1111/hex.13548

Wan, X., Stillman, F., Liu, H., Spires, M., Dai, Z., Tamplin, S., Hu, D., Samet, J. M., & Yang, G. (2013). Development of policy performance indicators to assess the implementation of protection from exposure to secondhand smoke in China. *Tobacco Control*, *22*(suppl 2), ii9–ii15. https://doi.org/10.1136/tobaccocontrol-2012-050890

Weiss, I., Stepanovic, S., Chinyemba, U., Bateman, J., Hemminger, C., & Burrows, E. (2016). Use of a Nutrition Behavior Change Counseling Tool: Lessons from a Rapid Qualitative Assessment in Eastern Zambia. *Frontiers in Public Health*, *4*, 179. https://doi.org/10.3389/fpubh.2016.00179

Williams, K. J., Gail Bray, P., Shapiro-Mendoza, C. K., Reisz, I., & Peranteau, J. (2009). Modeling the principles of community-based participatory research in a community health assessment conducted by a health foundation. *Health Promotion Practice*, *10*(1), 67–75. https://doi.org/10.1177/1524839906294419

Wilunda, C., Quaglio, G., Putoto, G., Lochoro, P., Dall'Oglio, G., Manenti, F., Atzori, A., Lochiam, R. M., Takahashi, R., Mukundwa, A., & Oyerinde, K. (2014). A qualitative study on barriers to utilisation of institutional delivery services in Moroto and Napak districts, Uganda: implications for programming. *BMC Pregnancy and Childbirth*, *14*, 259. https://doi.org/10.1186/1471-2393-14-259

Wright, T. B., Adams, K., Church, V. L., Ferraro, M., Ragland, S., Sayers, A., Tallett, S., Lovejoy, T., Ash, J., Holahan, P. J., & Lesselroth, B. J. (2017). Implementation of a Medication Reconciliation Assistive Technology: A Qualitative Analysis. *AMIA ... Annual Symposium Proceedings. AMIA Symposium*, *2017*, 1802–1811.

Yap, L., Wu, Z., Liu, W., Ming, Z., & Liang, S. (2002). A rapid assessment and its implications for a needle social marketing intervention among injecting drug users in China. *International Journal of Drug Policy*, *13*(1), 57–68. https://doi.org/10.1016/S0955-3959(01)00118-9

Zhou, W., Yu, Y., Zhao, X., Xiao, S., & Chen, L. (2019). Evaluating China's mental health policy on local-level promotion and implementation: a case study of Liuyang Municipality. *BMC Public Health*, *19*(1), 24. https://doi.org/10.1186/s12889-018-6315-7
